# A multivariate cell-based assay for blood-based diagnostics enhances lung cancer risk stratification

**DOI:** 10.1101/2024.11.04.24316585

**Authors:** Jason D. Berndt, Fergal J. Duffy, Mark D. D’Ascenzo, Leslie R. Miller, Yijun Qi, G. Adam Whitney, Samuel A. Danziger, Anil Vachani, Pierre P. Massion, Stephen A. Deppen, Robert J. Lipshutz, John D. Aitchison, Jennifer J. Smith

## Abstract

The indicator cell assay platform (iCAP) is a tool for blood-based diagnostics that addresses the low signal-to-noise ratio of blood biomarkers by using cells as biosensors. The assay exposes small volumes of patient serum to standardized cells in culture and classifies disease by machine learning analysis of the gene expression readout from the cells. We developed the lung cancer iCAP (LC-iCAP) as a rule-out test for nodule management in computed tomography (CT)-based lung-cancer screening. We performed analytical optimization, rigorous reproducibility testing, and assessed performance in a study with prospective-specimen-collection, retrospective-blinded-evaluation (PRoBE) design. LC-iCAP achieved an AUC of 0.64 (95% CI, 0.51-0.76) on the ROC curve in validation. Post-validation integration of the assay readout with CT-based features showed improved clinical utility compared to the Mayo Clinic model, with 90% sensitivity, 64% specificity, and 95% negative predictive value at 25% prevalence. The lung-cancer specific readout was enriched for hypoxia-responsive genes and was reproducible across different indicator cell lineages. This is the first validation study of an iCAP and the first application for early cancer detection. The LC-iCAP uses immortalized cells, is scalable and cost-effective and has a multivariate readout. This study supports its potential as a next-generation multivalent platform for precision medicine applications in multi-cancer screening and drug development.

**Key Points:**

- We developed the LC-iCAP, novel approach for liquid biopsies that uses cultured cells as biosensors. The cells detect cancer signals in serum and transduce them into standardized gene expression profiles, which are analyzed by machine learning for disease classification. The assay is inexpensive and scalable and has a multivariate readout with potential utility for precision medicine and multi-cancer early detection.
- A LC-iCAP-based lung cancer risk classifier demonstrated improved specificity compared to existing tests, suggesting meaningful clinical utility for managing indeterminate pulmonary nodules.
- We identified a lung-cancer specific transcriptional response to hypoxia in the assay readout, implicating HIF1A and HIF2A activity in the response consistent with known lung cancer biology and highlighting the platform’s mechanistic relevance.
- Standardized controls and validation studies demonstrated assay reproducibility, lineage stability, and detection of technical errors—supporting the platform’s readiness for clinical deployment.

## Introduction

The indicator cell assay platform (iCAP)^1^ is a novel approach that aims to overcome the low signal-to-noise ratio associated with direct measurement of blood biomarkers by using cultured cells as biosensors. Developing an iCAP involves exposing standardized, cultured cells to small volumes of serum from case and control participants, measuring a global differential gene expression response of the cells, and using machine learning to identify a subset of features for disease classification (Fig. 1*A*). This strategy takes advantage of the natural signal amplification and integrative sensing capacity of living cells, broadening the detectable analyte space while delivering a concise, multivariate readout. For deployment, the cell-based assay can be implemented in 96-well workflow using targeted high throughput platforms such as NanoString® or QuantiGene®.

**Figure 1.**
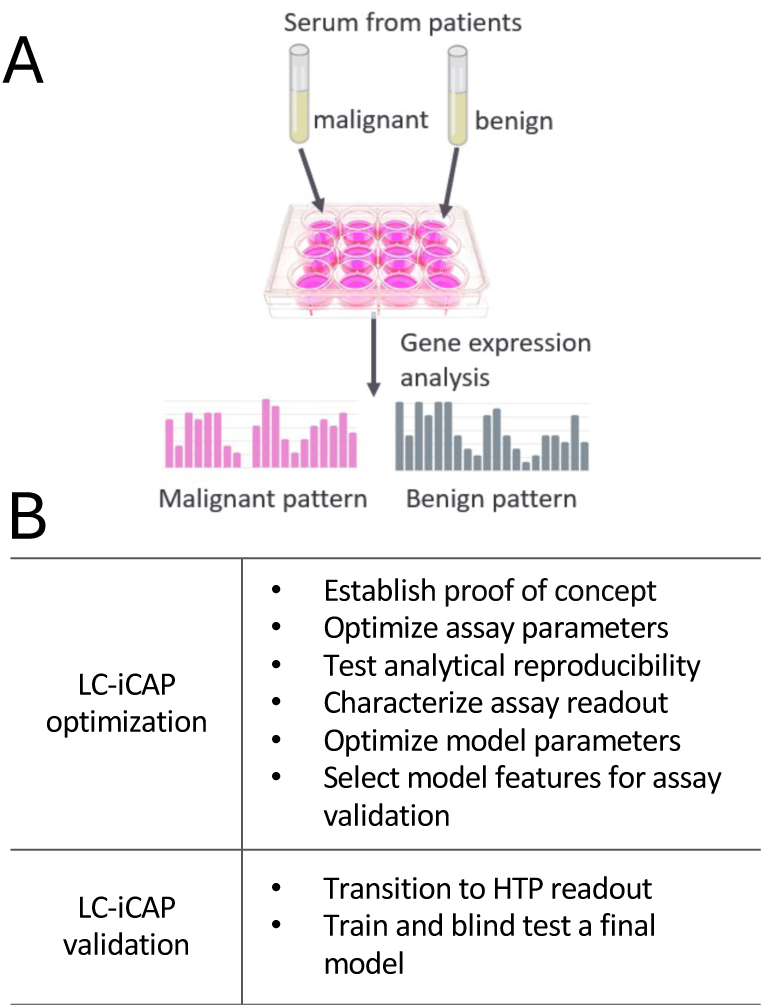
*A*, The iCAP for blood-based diagnostics. Standardized cells are exposed to serum from patients. Gene expression readout of the cells is used to develop machine learning models to predict disease. *B,* Steps of LC-iCAP development separated into two studies. *HTP*, high throughput.

Here, we describe the development of LC-iCAP, an iCAP-based assay for early detection of non-small cell lung cancer (NSCLC) in patients with pulmonary nodules identified by low-dose computed tomography (CT). The LC-iCAP was trained on serum from current and former smokers with indeterminate pulmonary nodules (IPNs) and validated using a retrospective-blind-temporal design. In validation, the assay achieved an AUC of 0.64 (95% CI, 0.51–0.76). After validation, when integrated with CT data, LC-iCAP yielded 90% sensitivity, 64% specificity, and a 95% negative predictive value (NPV), assuming a 25% disease prevalence in the intended use population.^2,3^ These results represent improved performance over existing blood-based rule-out tests and suggest the potential for LC-iCAP to provide actionable results for a greater proportion of patients.

Indeterminate pulmonary nodules present a significant clinical challenge: under current guidelines, patients with malignancy risk below 5% or above 65% have clear treatment pathways, but the majority fall into an intermediate-risk category (5–65%) where management is uncertain. Many of these patients undergo invasive and costly diagnostic procedures despite ultimately having benign disease.^4,5^ Non-invasive rule-out type tests are needed to identify those with low risk of cancer to save those with benign nodules from invasive and expensive testing.^6,7^ Such a test must have an NPV of at least 95% to reclassify cancer risk to less than 5% to reduce unnecessary interventions and improve clinical outcomes by shifting treatment plans.^8^ While two blood-based rule-out tests are commercially available,^9,10^ their limited standalone performance and low specificity when combined with CT data, restrict their utility. A third test outperforms these blood tests but requires invasive bronchoscopy.^11^ LC-iCAP offers a technically distinct and clinically valuable solution by providing measurable performance independent of imaging and enhancing classification accuracy when integrated. As such, it may increase physician confidence, reduce unnecessary procedures, and improve outcomes for patients undergoing lung cancer screening.

The iCAP is a multivalent, scalable platform that is orthogonal to existing diagnostic approaches and supports high-throughput implementation. Its flexible architecture warrants further development for broader applications in precision medicine, including multi-cancer early detection (MCED), combinatorial diagnostics, and disease stratification.

## Methods

The study design description below follows recommendations of TRIPOD (Transparent Reporting of a multivariable prediction model for Individual Prognosis Or Diagnosis).^12^

### Participants and specimen characteristics

The study used archived serum samples collected from adult patients from non-vulnerable populations performed with IRB approval (WCG IRB study 1283522). Banked specimens and clinical data used in this study were from subjects enrolled in the following previously IRB-approved studies: “Molecular Predictors of Lung Cancer behavior,” (NCT00898313, Vanderbilt University), “Gene-Environment Interactions in Lung Cancer” (IRB 806390, University of Pennsylvania), and “A Case Control Study of Smokers and Non-Smokers” (IRB 800924, University of Pennsylvania). The study consent forms had provisions allowing use of their samples for future research purposes. Patient identifiable information was not provided to the research team and was not used in this study. All identification numbers used in the manuscript are not known to anyone outside the research group.

Patient attributes and serum characteristics are shown in Table I and Data Files 1 and 4. All samples were collected within 3 months of the CT scan (and diagnostic biopsy, if performed) and prior to any invasive procedure, including surgical lung biopsy. All malignant nodules were diagnosed by follow-up pathological diagnosis; all were non-small cell lung cancer (NSCLC) except for 3 small cell lung cancers. 72% of malignant nodules in the blind test set were stage I and 22% were stage II. Benign nodules were diagnosed by either biopsy with a definitive benign histological diagnosis or by 2 or more years of follow-up with serial imaging. All patients had no known other cancers at time of screening and no previous cancer in the five years preceding the blood draw excluding previous skin cancers treated with surgery only (no radiation or chemotherapy). Serum samples were collected using the protocol recommended by the early detection research network (EDRN),^13^ stored at -80°C or below after collection, and unless otherwise stated, thawed once prior to LC-iCAP analysis.

**Table 1.** Samples used for LC-iCAP optimization and validation. Patients had IPNs identified by CT scan that were classed as malignant or benign by either diagnostic biopsy or resection or by 2 or more years of serial imaging. Patents had no other known cancer at the time of CT scan. Cohorts 1-4 were from Vanderbilt University. A 5th cohort from University of Pennsylvania was used for sample quality analysis only (see Methods). LC-iCAP optimization used cohorts 1-3. LC-iCAP validation used cohorts 1-3 for model training and cohort 4 for blind validation. Cohort 4 was temporally independent as it was collected 9 years later and assayed 1 year later than the other samples. *IPN*, indeterminate pulmonary nodule; CT, low-dose computed tomography.

| Class | Description | cohort 1 | cohort 2 | cohort 3 | cohort 4<br>BLIND |
| --- | --- | --- | --- | --- | --- |
| Serum from patients with non-calcified IPNs identified by CT scan between 5-30 mm in diameter (95% > 7 mm) | Case: patients with malignant nodules | 6 | 53 | 50 | 40 |
|  | Control: patients with benign nodules | 6 | 50 | 50 | 40 |
| subtotal |  | 12 | 103 | 100 | 80 |
| Total sample count |  | 295 |  |  |  |

### Study design

The study was an observational case versus control study with blind validation following prospective-specimen-collection, retrospective-blinded-evaluation (PRoBE) design.^14^ Serum samples were collected at the time of CT scan from patients representative of the target population with the intention of using the serum for developing liquid biopsies in the future. After outcome status was ascertained by follow-up testing, case and control subjects were selected randomly from the biobank and their serum was used for developing the LC-iCAP.

LC-iCAP development included two stages, assay optimization and assay validation (Fig. 1*B*). Assay optimization included rigorous analytical validation and intermediate modelling studies, first to establish proof of concept of the assay and then to optimize the parameters and hyperparameters of the models. This involved dividing samples into training and held-out validation sets, for tuning model parameters and hyperparameters and estimating model performances while training. Assay validation involved first training while estimating model performances using nested cross validation, and then testing fully specified models using a blind test set. During assay development, sample and data quality were evaluated and in the final model configuration, data from samples with technical assay failures and low-quality samples were excluded.

The study used a total 528 serum samples from two sources, Vanderbilt University and University of Pennsylvania (Table I), including a blind cohort for final assay validation (*shaded*). The blind ‘test set’ was collected 9 years later and analyzed 1 year later than the other cohorts for temporal independence. For intermediate modeling, the sample sizes were selected based on other liquid biopsy studies at similar stages of development.^9,15^ For blind testing, the sample size was selected to have power to detect significant performance of a model with AUC of ROC >= 0.62.^16^

Sample processing batch structure is described in Data files 1 and 4. Each cohort was shipped separately and assayed by LC-iCAP in batches of 20-50 samples. Each experimental batch had ∼1:1 ratio of case and control samples, which were roughly balanced for patient gender, age and smoking history. A pair of standard controls were included on each 12-well plate for quality control (QC) consisting of either technical replicates of a reference serum sample from an unaffected male patient (cohorts 1-2), a pair of case and control pooled serum controls (cohort 3), or a pair of DMOG and PBS chemical controls (cohorts 4-5). LC-iCAP gene expression was measured by RNA-seq for cohorts 1-3 for intermediate modeling, and by NanoString Plexset for all cohorts for final assay validation. For cohort 5, only the DMOG samples were included in the NanoString Plexset analysis, excluding the PBS controls, to fit all samples on a single plate.

### Quantification of hemolysis of serum samples

Prior to LC-iCAP analysis, thawed patient serum was evaluated for the breakdown of red blood cells (hemolysis), which has known interference with clinical biochemical tests.^17^ Blind samples were visually compared to a reference card^18^ by a first scientist and given a score on a gradient of increasing hemolysis from 1-7. In addition, samples were photographed with an iPhone on a white background and later evaluated by a second scientist using the same approach. Sample ratings were averaged and round to the nearest integer. For all modeling conducted after the pilot study, all samples with average scores greater than 3 (> 50 mg/dL of hemoglobin) were omitted from the study. Hemolysis scores are shown in Data files 1 and 4.

### Analytical parameters of LC-iCAP assay

Unless otherwise indicated, the following standard protocol was used: 2 x 10^6^ lung epithelial cell indicator cells (16HBE14o- (16HBE)) were thawed and plated in a T75 flask in RPMI with 10% FBS (complete medium). After 2 d cells were dissociated with 0.25% trypsin-EDTA for 10 min at 37 °C and plated at 30,000 cells/cm^2^ in 12-well Eppendorf moat plates in complete medium. After 24 h, cells were rinsed once with RPMI and incubated for 24 h with in 1 mL RPMI with 5% patient serum. Media was removed, lysis buffer was added, and cells were stored at -80°C for up to two weeks before RNA isolation. For cohorts 1 and 2 used in assay optimization, total RNA was isolated manually using RNeasy Mini Kit (Qiagen), for cohorts 3-5, RNA isolation was automated using a MagMax mirVana kit (Invitrogen A27828) on either a KingFisher Flex or a Kingfisher Duo Prime as per the manufacturer’s recommendation. RNA was eluted in 100 µL of elution buffer, quantified using a Qubit (Qubit RNA BR Assay Kit), and stored at -80°C. Transcript abundance levels were quantified using either RNA-Seq or NanoString as described below. For cohort 3, cells were passaged an additional time before initiating the experiment.

### LC-iCAP proof-of-concept study

To establish assay feasibility, LC-iCAP RNA-seq data for cohort 1 was used to train a model for lung cancer detection and tested on cohort 2. To identify model features, raw counts from cohort 1 data were used for differential expression analysis and identified 239 differentially expressed genes (DEGs) (Benjamini-Hochberg false discovery rate (FDR) < 0.05). Next, to develop the LC-iCAP classifier, all RNA-seq count data from cohorts 1 and 2 were normalized using the DESeq2 rlog transformation and a series of random forest classifiers (R: randomForest package) were parameterized on cohort 1 using increasing numbers of DEGs in increasing order of FDR as features (5, 10, 20, 25, 50, 75, 100). The 103 samples of cohort 2 were used to test the model performance.

For hierarchical clustering of LC-iCAP data from cohorts 1 and 2, clustering was performed based on the expression of the top 20 differentially expressed in LC-iCAP RNA-seq data from cohort 1. The analysis utilized DEseq2 rlog-transformed counts from the LC-iCAP RNA-seq data, normalized to the mean expression of the benign samples of the same iCAP experimental batch and gene rank was based on median absolute deviation. Three outlier samples were identified and removed (all from the benign class).

### RNA-Seq analysis of LC-iCAP RNA

100-650 ng of total RNA per sample was used for automated library preparation and RNA sequencing (RNA-seq) performed by either Covance (cohorts 1-2) or Azenta Life Sciences (formerly Genewiz) (cohort 3). Strand-specific library prep was performed with PolyA selection using TruSeq RNA library Prep Kit (Illumina) with unique dual indices (IDT) and resulting DNA was sequenced on a HiSeq 4000 (Illumina) with paired end 150 bp reads. RNA-seq data were processed using a custom workflow including adapter read trimming using *trimmomatic*,^19^ genome reference alignment to HG37 (pilot study) or HG38 using *STAR*;^20^ and gene-level transcript quantification using R-featureCounts.^21^ ERCC spike-ins and genes with mean absolute counts < 10 were removed. Quality was assessed using MultiQC, dupRadar and GATK estimate of library complexity. Where indicated, counts were adjusted to correct GC bias using the FQN^22^ or CQN^23^ determined by the R-EdgeR package^24^. Differential expression analysis was done using R-DESeq2.^25^ GSEA analysis was done using the R-fgsea package in combination with the 50 Hallmark pathway modules from MsigDB.^26^

For proof of concept analyses, read duplicates were removed by using –ignoreDups setting in R:featureCounts. For all other assay optimization analyses, the data were normalized for heteroskedasticity by variance stabilizing transformation (VST) using the R-DESeq2 package and for inter-iCAP batch variation using removeBatchEffect from the R-limma package.^27^ Outlier samples were identified using robust principal components analysis (ROBPCA) implemented in the R-rrcov package.^28^

### Development of LC-iCAP standard controls

Pooled serum standards were prepared, consisting of technical replicates of pooled patient serum for each case and control class. Each pool consisted of a mix of serum from 8 different subjects selected from cohorts 1 and 2 based on availability and class separation in hierarchical clustering analysis of LC-iCAP-RNA-seq data (Fig. 2*B*). Each serum pool was made by thawing aliquots of serum from each of 8 subjects, pooling, mixing, aliquoting the serum, and then flash freezing in liquid nitrogen before storing at -80°C. Dimethyloxalylglycine (DMOG) and PBS were used as a pair of chemical standards to monitor assay performance. To develop the control, we first characterized the response of indicator cells to 6 concentrations of DMOG (0, 0.025, 0.05, 0.1, 0.25, 0.5 mM; Cayman Chemical) by measuring the gene expression readout of the 74-gene development gene set (Data file 2A) using NanoString. Responsive genes were identified as those whose expression fit a linear model as a function of DMOG concentration (p-value < 0.05) (data not shown). DMOG was used at 0.25 mM for monitoring assay performance; for most responsive genes, this condition was in the linear range of the model and had a similar magnitude of responsiveness compared with patient serum. DMOG was resuspended at 50 mM in PBS, aliquoted and stored at -20°C and thawed on ice before use.

**Figure 2.**
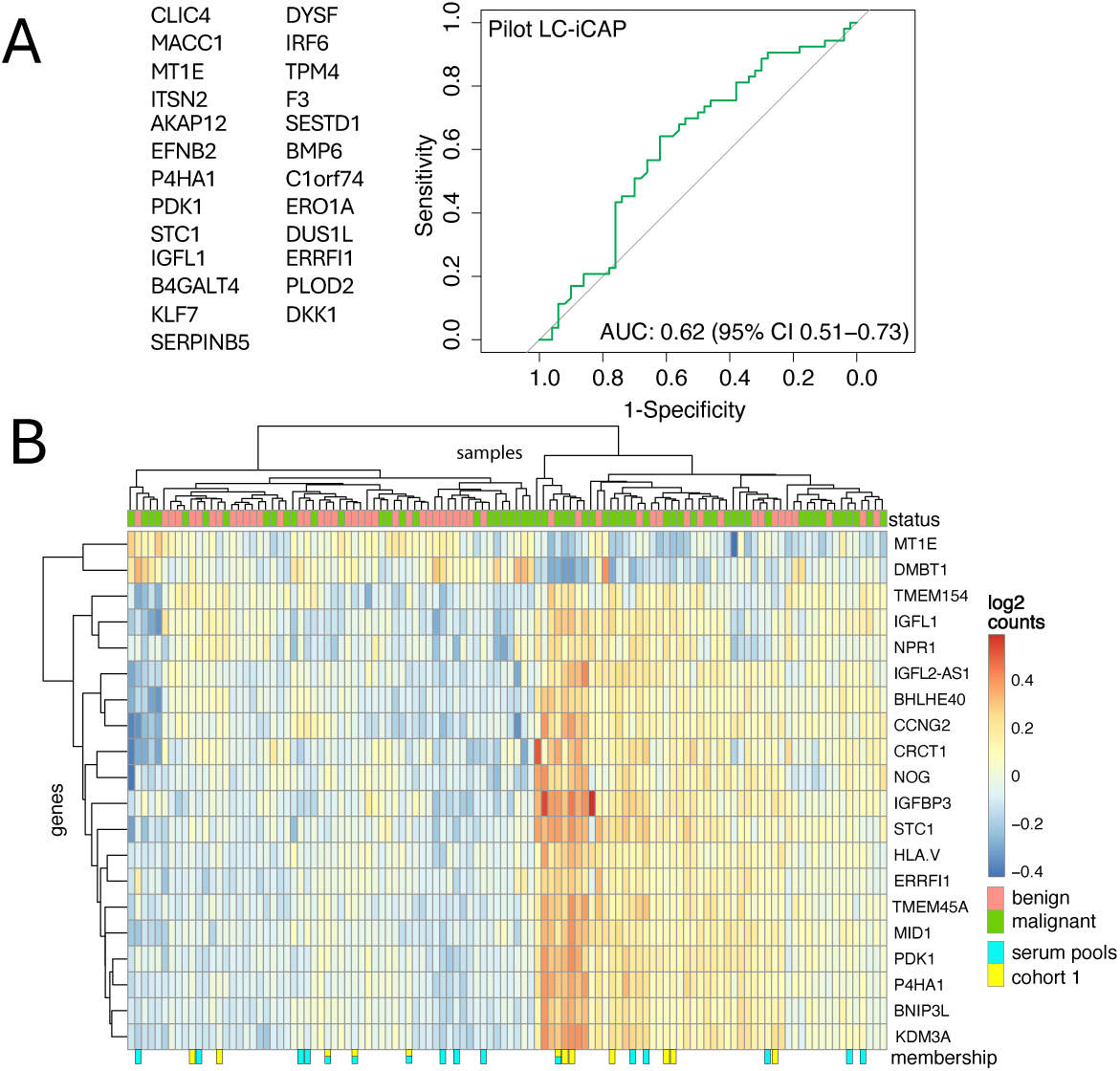
Proof of concept of LC-iCAP. ***A.*** ROC curve showing performance of a pilot LC-iCAP model trained on cohort 1 (n=12) and tested on held-out cohort 2 (n = 103) using 25 gene expression features (*left*). ***B.*** Hierarchical clustering of cohorts 1 and 2 based on LC-iCAP gene expression shows grouping of samples by class. Clustering was based on the top 20 differentially expressed genes in cohort 1 based on median absolute deviation. Dendograms show two distinct sample clusters broadly separating case and control samples (*top*). 3 outlier samples were omitted. Samples used to make the biological standards (serum pools) and the 12 samples of cohort 1 are marked in cyan and yellow, respectively (*bottom*).

### Optimization of LC-iCAP experimental parameters and LC-iCAP reproducibility testing

The pooled serum standards described above were used for QC and for reproducibility and optimization studies. 4 technical replicates of each case and control serum pool were analyzed for each LC-iCAP parameter and the number of significantly DEGs (FDR <0.1) for each configuration were compared. Differential expression was measured by analysis of the LC-iCAP development gene panel using NanoString nCounter® technology and/or by RNA-seq using HiSeq4000^TM^ (Illumina). Configurations tested were various serum concentrations (1%, 5%, 10%, and 20%), serum incubation times (6 hours, 24 hours), and 4 cell types (16HBE, A549, MRC5, and Nuli-1). Reproducibility testing included comparison across 3 different 16HBE expansion batches and across different days, and different detection platforms.

### Western blot analysis

Samples were processed in the LC-iCAP using standard parameters and protein was isolated and quantified using a BCA kit (Pierce). Equal protein for each sample were loaded on 12- or 15-well NuPage 4-12% Bis-Tris gels with Thermo PageRuler Plus prestained protein ladder and analyzed by western blotting according to LICOR Odyssey recommendations using PVDF membrane (Immobilon-FL) and probing with rabbit anti-HIF1a (D157W) XP^®^ (or rabbit mAB HIF-2a (D9E3)) and mouse anti-beta-actin primary antibodies followed by LICOR NIR secondary antibodies. All primary antibodies were from Cell Signaling Technologies. Membranes were scanned on a LICOR Clx Odyssey imaging system, proteins were quantified using LiCOR Image Studio Lite and data analysis was done in Excel. Western blot images are shown in Figure S5.

### Sample and Gene Filters for Comprehensive Modeling Study in Figure 5

Data from 165 samples across cohorts 1-3 were partitioned into training and validation sets, and quality filters were applied, resulting in 137 samples for modeling (Table I, Data file 1). These data were used for a comprehensive modeling study including 13 different feature selection methods x 8 combinations of 3 optional filters. The three sample filters were: 1) samples with predicted forced expiry volume (FEV) < 50% (based on data showing that low lung function affects LC-iCAP readout (Fig. S6)), 2) samples from never-smokers, and 3) samples from a low-quality RNA-seq batch (identified by QC analysis with assay standards; Fig. S3). The 13 feature selection methods are described in Data file 3 and are based on 3 approaches to identify case versus control differential expression in the training set: 1) Analysis of DEGs across samples from all batches, 2) analysis of DEGs within each individual experimental batch, and 3) GSEA of individual experimental batches.

### Gene sets for targeted analysis by NanoString

Gene-specific capture probes for NanoString nCounter analysis were synthesized by Integrated DNA Technologies (IDT). The NanoString development gene set was used for LC-iCAP parameter optimization and reproducibility studies. The set consisted of 96 genes including 74 features with case versus control differential expression in cohort 1 selected for detecting the LC-iCAP readout and 7 housekeeping genes for normalization (Data file 2A). The NanoString deployment gene set was used for training and testing the final LC-iCAP models in the assay validation study using a NanoString Plexset readout. The gene set consisted of 95 genes including 85 candidate features for modeling (66 selected in the model optimization (Fig. 5) and 19 of the 25 genes from the initial proof of concept model not already in the list), 1 control gene responsive to DMOG and 9 housekeeping genes for normalization (Data file 3D).

### NanoString Plexset^™^ analysis of LC-iCAP RNA

Gene expression analysis of total RNA samples from the LC-iCAP was performed using NanoString Plexet^™^ technology, a direct detection approach for multiplexed gene expression analysis of up to 96 samples per run with no PCR amplification. For this analysis, the ‘deployment gene set’ was used consisting of 85 target genes and 9 candidate housekeeping genes (Data file 3). Data were analyzed using an nCounter® Analysis System by the Genomics Resources Center at Fred Hutchinson Cancer Research Center following manufacturers recommendations. First, probe hybridization was performed in solution where gene-specific capture probes and reporter probes attached to fluorescent barcodes (code sets) were used for detection of each of 96 transcripts in each sample (see Data file 3). Each reaction included LC-iCAP RNA from patient samples (140 ng) or from DMOG or PBS controls (100 ng), amounts that were pre-optimized in a calibration experiment to avoid saturation. Next, samples were pooled in groups of 8 and loaded onto an nCounter Prep Station for automated excess probe removal and binding of the probe-target complexes on the surface of the cartridge by streptavidin-biotin linkage to the capture probes. Cartridges were placed in the nCounter Digital Analyzer for data collection, where molecules of RNA were counted by using the target specific “color codes” generated by a string of six fluorescent spots on each reporter probes.

LC-iCAP analysis was done using our standard approach except serum samples for cohorts 1-2 were thawed twice before analysis. LC-iCAP RNA samples were processed on 7 plates with up to 96 samples per plate (Data file 4). Each plate included 8 positive controls composed of *in vitro* transcribed RNA transcripts and corresponding probes, and eight negative controls consisting of probes with no sequence homology to human RNA for lower limit of detection analysis. One positive and one negative control were used for each of the 8 multiplexed samples in each lane. Analysis of training set samples (plates 1-6) and blind test set samples (7^th^ plate) were separated by one year using two different code set manufacturing lots. To facilitate code set lot normalization in downstream processing, technical iCAP replicates of a calibration sample were run with code set lot 1 (training samples) and code set lot 2 (blind test samples).

Processing of raw Plexset data was performed using nSolver software following manufacturer’s recommendations (MAN-C0019-08) as specified below. Data points below the lower limit of detection were floored to a threshold of 20 counts. Next correction for technical variation was done by housekeeping (HK) gene normalization, for which the expression value of each gene was divided by the geometric mean of 5 stably expressed housekeeping genes (ABCF1, FCF1, GUSB, POLR2A, and SDHA) in the same sample. Next, within-plate code set calibration was performed to normalize the 8 different sets of barcodes used for each of the 8 rows. Finally, code set lot calibration was done to normalize lot 1 (training samples in plates 1-6) and lot 2 (blind test samples in plate 7) using the “batch normalization” tool of nSolver. Prior to using Nanostring data for modeling, raw counts were log2 transformed, and outlier removal was performed using ROBPCA^29^ analysis of housekeeping genes only.

### NanoString Plexset reproducibility

To investigate the effectiveness of the plate normalization (within training samples (plates 1-6)) and code set lot calibration (between training and blind test samples), hierarchical clustering was performed on the chemical and biological controls in the LC-iCAP Plexset data. While good plate to plate reproducibility was observed, code set lot mis-calibration was identified which resulted in differential expression of in the LC-iCAP between control samples of the training and test sets (p-value < 0.005) (compare lot 1 versus lot 2 in Fig. S9, Data file 4).

### Modeling

GLM models were implemented in the R-glmnet package.^30^ RF models were implemented in the R-caret package^31^ using mtry values that were automatically selected for each seed using the default settings. For the proof-of-concept model, RF models were trained by leave-one-out cross validation with 10 random seeds and tested on a held-out independent validation set. For the large-scale modeling study in Figure 5, RF models were training set by leave-one-out cross validation with 20 random seeds and 50 resampling iterations and tested on a held-out independent validation set. Each model had a maximum 20 gene features selected based on variable importance score and all models were repeated with downsampling of the training set to balance the number case and control samples within each experimental batch to assess model robustness. Top models were selected based on performance on the validation set (using either AUC or specificity at a fixed sensitivity of ≥ 95%).

For modeling in the final assay validation study with LC-iCAP NanoString Plexset data, RF or GLM models were trained and validated using 5-fold nested cross-validation with 50 subsamples per seed, implemented using R-nestedcv framework.^32^ Cross-validation was repeated 10 times with different random seeds, and RF models used 500 trees. Features were log2 normalized iCAP gene expression values, with inclusion of smoking status as a covariate. When applied, feature selection was performed using Elastic Net regularization of R-glmnet, with the optimal alpha (mixing parameter) selected by cross-validation across values from 0.1 to 1. Stable features were defined as those consistently selected across 50 subsamples, each stratified by outcome and smoking status. Optimal modeling conditions were selected based on outer test set AUC performance, and seeds used for blind testing corresponded to the 50th or 75th percentile of AUCs observed across training repetitions.

### Blind testing

For blind testing, 80 serum samples were selected by Vanderbilt University to match the samples used for model development with two additional criteria: The selection only included samples of high quality (those with an estimated hemolysis less than 50 mg/dL and storage times of 4 years or less) and samples from patients who were current and former smokers.

The sample size of 80 was selected to have power to identify significant models with AUC of ROC ≥ 0.62 using an established method.^16^ Patient status and clinical data were blind to researchers at time of prediction. However, information on approximate overall ratio of case to control samples, and nodule size and smoking status were available. Samples were shipped in one batch and processed in random order in 4 LC-iCAP batches and one Plexset plate using a new lot of code sets. LC-iCAP NanoString Plexset processing included 8 DMOG controls for quality control (QC) analysis. Using these controls, we identified and omitted genes with significant differential expression between DMOG controls of code set 1 (training samples) versus code set 2 (test samples) (p-value < 0.005). 9 models using iCAP features were tested on the blind test set, including 5 before and 4 after quality control-based gene omission.

### LC-iCAP models M3 and M4

RF models M3 and M4 selected for blind testing used either all 35 Plexset genes remaining after code set lot QC analysis or a subset of 14 genes remaining after feature selection, and both used smoking status as a covariate.

### Mayo Clinic Model

Pre-test risk of cancer malignancy was calculated for the test set by researchers at Vanderbilt University using Solitary Pulmonary Nodule (SPN) Malignancy Risk Score (Mayo Clinic model).^33^ This model uses 6 clinical risk factors including nodule spiculation, upper lobe location, smoking status (current or former vs non), nodule diameter, age, extrathoracic cancer diagnosis ≥ 5 years prior. The test excludes patients with prior lung cancer diagnosis or with history of extrathoracic cancer diagnosed within 5 years of nodule presentation.

### Integrated LC-iCAP model

Prototype integrated classifiers were developed after blind testing by integrating LC-iCAP model M3 or M4 with the Mayo Clinic model using an approach similar to that used for the Nodify XL2.^10^ This approach is a decision tree model whereby the Mayo model’s output is conditionally adjusted by a fixed amount based on a threshold established using the liquid biopsy readout: For LC-iCAP probabilities below the threshold, the Mayo model’s output was reduced by a fixed amount and for probabilities above the threshold, the Mayo model’s output was used without adjustment. See Figure S12 for the technical parameters.

The integrated model performance on the blind test set was compared to that of the Mayo model by generating ROC curves for both models and comparing fixed decision thresholds corresponding to maximum specificity at ≥ 90% sensitivity and ≥ 95% NPV (with 25% disease prevalence) using exact binomial version of McNemar’s test.^34^ Model calibration was not required for this comparison because it used points from the ROC curves corresponding to specific performance metrics rather than the absolute probability estimates. For verification, both models were calibrated to the test set prevalence using logistic regression (R-caret), and identical ROC curves were regenerated.

### Control for error and bias

Key biological resources were authenticated: an aliquot of the 16HBE indicator cells was validated midway through the study externally at IDEXX bioanalytics by CellCheck 16^TM^ Plus, and it passed all three tests. The cell line of origin was confirmed to be correct with 15 of 16 markers of the 16STR profile matching 16HBE from Sigma and no contamination from *mycoplasma spp.* or other species was detected. Serum samples were assayed for hemolysis and storage time and omitted if above established thresholds. Patient gender age and smoking history were approximately balanced between classes. All DNA constructs were sequenced. All chemical resources were from reputable commercial sources.

Controls in data generation included: blinding researchers to disease status; randomizing sample positions; balancing classes within batches; balancing patient attributes between batches; using moat plates to control edge effects in the cell-based assay; developing standard controls and using them to monitor assay performance and batch effects; performing reproducibility studies; measuring RNA integrity before RNA-seq and omitting samples below a threshold 7; and detecting and correcting biases from RNA-seq (GC bias and batch effect) and NanoString Plexset analyses (batch effect).

Modeling practices were used that control for biases and overfitting including: training sets were balanced with approximately equal numbers of case and control samples; all models contained fewer features than samples to prevent overfitting (except for the pilot model, which had 25 features and 12 samples); validation was conducted using independent samples or nested cross-validation, both robust methods that mitigate overestimation of model performance regardless of sample size;^35^ different gene expression detection methods were used for feature selection and final model development to avoid effects of platform biases on model performance; final models were evaluated by blind testing with independent samples; and samples for blind testing had temporal independence from the training set for both sample collection and processing, increasing the rigor of the test.

## Results

The iCAP is a multivalent platform for blood-based diagnostics that captures complex, disease signals from patient serum by using standardized cells as biosensors. Multi-variate gene expression responses of the cells are then analyzed using machine learning to predict cancer status (Fig. 1*A*). To evaluate the utility of the iCAP for lung cancer screening, we optimized and validated the LC-iCAP for classification of indeterminate pulmonary nodules identified by CT scan. The assay was developed in 2 stages outlined in Figure 1*B*.

### Study Summary

The study had PRoBE design, with prospective-specimen-collection and retrospective-blinded-evaluation of a disease classifier.^14^ Serum samples were collected from patients with IPNs identified by CT scan and later characterized as malignant or benign based on diagnostic biopsy/resection or ≥2 years of serial imaging follow-up (Table I).

Archived samples were used to develop the LC-iCAP in two stages: assay optimization and assay validation (Fig 1B). Optimization included establishing standardized controls, assessing assay reproducibility, and identifying sources of preanalytical and analytical variation. It also included optimization of model parameters and selection of LC-iCAP gene expression features for final model development in the next stage. Classifier development during this stage used training and held-out validation sets to evaluate model performance and guide parameter and hyperparameter tuning.

Next, a set of 85 candidate features selected during assay optimization were used to analyze patient samples in a high-through put version of the LC-iCAP for final model development and validation. This second stage used nested cross-validation for model training, followed by blind testing on a temporally independent test set to assess the performance of fully specified models. Sample sizes were guided by power calculations, and sample exclusions were made before validation. Validation sets were kept fully independent from tuning sets, and the research team remained blinded to test set classes until all predictions were finalized. To minimize overfitting, models were constrained to use fewer features than samples.^36,37^ Additionally, feature selection and final model testing were performed on different gene expression platforms (RNA-seq and NanoString PlexSet, respectively) to reduce analytical bias. Detailed methods follow below.

### LC-iCAP Optimization

#### Proof of Concept

To establish proof of concept for the LC-iCAP assay, we trained and tested an initial classification model. We first acquired Cohort 1 from Vanderbilt University consisting of serum samples from patients with IPNs, including six with lung cancer and six with benign nodules (Table I, Data File 1). Samples were processed in the LC-iCAP assay alongside reference serum controls. RNA-seq analysis identified 239 differentially expressed genes (DEGs) between cases and controls (FDR < 0.05, Benjamini-Hochberg) and functional enrichment analysis using STRING revealed that these DEGs formed a significantly interconnected network with enrichment for HIF1A signaling (KEGG) and response to hypoxia (Gene Ontology), both of which are biologically relevant to lung cancer.^38^ We next trained a Random Forest (RF) classifier on Cohort 1 data using the top 25 DEGs ranked by FDR as features with 10 random seeds and internal cross-validation to generate a pilot LC-iCAP model for lung cancer prediction.

We tested the pilot model on LC-iCAP data from an independent set of 103 serum samples (Cohort 2; Table I, Data File 1). The classifier demonstrated significant performance with a median AUC of 0.62 (95% CI: 0.51–0.73; Fig. 2A). Similar performance was observed when models were trained using the top 50, 75, or 100 DEGs (data not shown).

To further evaluate signal generalizability, hierarchical clustering was performed on Cohorts 1 and 2 using the top 20 DEGs from Cohort 1. This approach broadly separated samples into two class-associated clusters (Fig. 2B), supporting the presence of lung cancer-specific signals in the readout.

Together, these data establish feasibility of the LC-iCAP approach for lung cancer detection. However, the results also suggest potential sources of variability in the readout, including patient heterogeneity, serum quality, and analytical factors inherent to the assay or RNA-seq. These sources of noise were investigated further during assay optimization.

#### Assay Controls and Standards

Biological and chemical standardized controls were developed to support assay optimization, monitor performance, and assess reproducibility.

Biological standards consisted of pooled case and control sera, each generated by combining aliquots from eight individuals in Cohorts 1 and 2 (indicated in Fig. 2B). These serum pools were assayed using the LC-iCAP with a targeted NanoString panel comprising 58 DEGs from Cohort 1, along with 30 additional genes (Fig. S2; Data File 2). NanoString analysis of four technical replicates of each pool identified 55 genes as differentially expressed between case and control samples (FDR < 0.1) (Fig. 3A), establishing a baseline biological signal for use in subsequent optimization studies.

**Figure 3.**
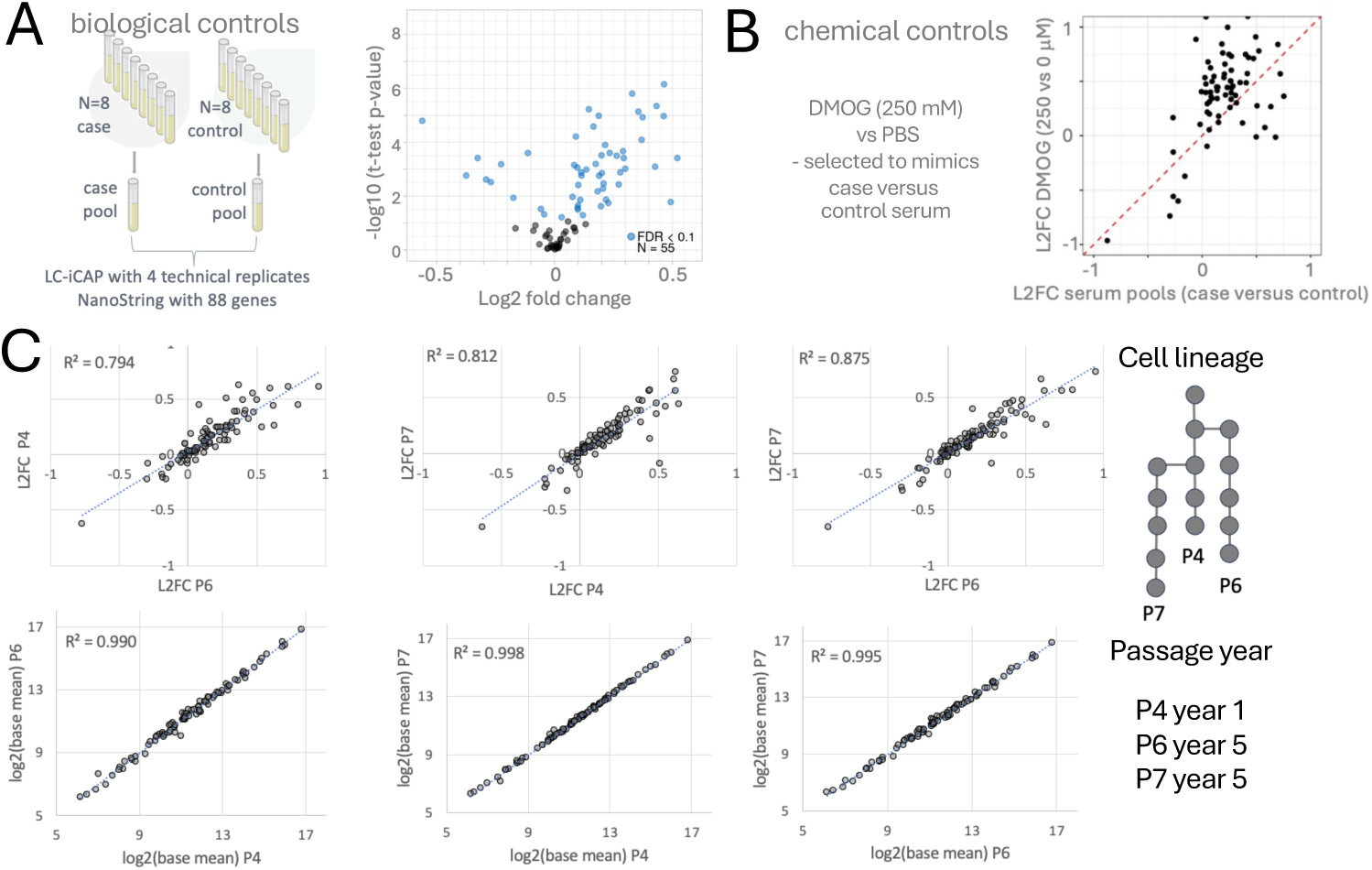
Development of assay controls and demonstration of stability of LC-iCAP indicator cells. ***A.*** Biological controls were pools of case and control patient serum samples. The volcano plot shows the quantitative assay readout for these controls - differential expression of ∼55 of 88 genes measured by NanoString. ***B.*** Chemical controls were DMOG versus PBS, which were selected to mimic the HIF1A/hypoxia response to case versus control sera of cohort 1. The scatter plot shows that the LC-iCAP-NanoString readout using the chemical and biological controls is broadly coherent. ***C.*** LC-iCAP differential expression is reproducible across 3 different passages of indicator cell generated over 4 years (P4, P6, and P7). LC-iCAP NanoString data were generated for 4 replicates of case and control serum pools across the three different passages of indicator cells. Graphs show pairwise comparisons of expression levels (*lower*) and differential expression levels (*upper*). The R² values of the linear models indicate that the three cell passages exhibit similar variability, supporting the stability of the cell line for clinical assay development. Other metrics of comparison are shown in Figure S2*A*. *L2FC*, log2 fold change.

Chemical standards included a phosphate-buffered saline (PBS) control versus DMOG, a small-molecule agonist of the hypoxia pathway, selected to mimic the malignant serum-induced hypoxia response observed in Cohort 1. In an initial experiment (see Methods), DMOG exposure in indicator cells triggered differential expression of 24 of the 58 development genes. A scatter plot of log-fold changes for both standards demonstrates strong concordance between the DMOG and serum-induced responses across the NanoString panel (Fig. 3B), supporting the use of both biological and chemical controls to benchmark assay behavior.

#### Optimization of Assay Parameters

Assay parameters were optimized with the goal of detecting and controlling for unwanted variation from analytical sources to improve stability and magnitude of differential expression and thus model performance. To do this, we assayed 4 technical replicates of case and control serum pools in the LC-iCAP with the NanoString readout under various assay parameters and optimal conditions were selected that maximized the number of significant DEGs and/or the separation of the classes by principal component analysis (PCA). We tested 4 candidate indicator cell types (16HBE, A549, MRC5, and Nuli-1); 2 serum incubation times (6 hours, 24 hours); 4 serum concentrations (1%, 5%, 10%, and 20%); and the effect of Trichostatin A (TCA) addition, an inhibitor of the hypoxic response. The optimal LC-iCAP parameters were found to be a 24 h incubation of either 5% or 10% serum with 16HBE lung epithelial indicator cells in the absence of TCA, which matched the baseline conditions used in proof of concept (Fig. S1). To avoid potential bias from measuring only a subset of genes, RNA samples from the TCA and cell type experiments were reanalyzed by RNA-seq, which yielded similar results (data not shown).

In addition to the analytical optimizations of the cell-based assay listed above, we detected and corrected sources of noise arising from LC-iCAP and RNA-seq batch effects, and artifactual duplicate reads and GC biases in the RNA-seq data. All corrections improved model performance and increased the detected number of DEGs across classes (data not shown), and thus were implemented in data processing going forward.

#### Analytical Reproducibility Testing

We next evaluated the analytical reproducibility of the LC-iCAP by testing gene expression stability across multiple experimental conditions. To address the long-term stability of the cells, we tested the serum pool standards in the LC-iCAP using three different lots of indicator cells derived from distinct lineages, expanded over a four-year span. Pair-wise comparisons of gene expression between lots showed strong linear correlations (R^2^ > 0.99; Fig 3*C*). Comparisons of differential expression also showed good agreement (R^2^ > 0.79; Fig 3*C*) and PCA plots demonstrated consistent separation of case versus control samples across all lots (Fig. S2*A*). These results support the long-term stability of the cells for clinical application.

Next, batch-to-batch and platform reproducibility were tested. We tested the pooled serum standards across different LC-iCAP batches run on different days, different gene expression detection platforms (RNA-seq versus NanoString) and different NanoString batches. Gene expression profiles remained highly correlated across all conditions (R^2^ > 0.99) and differential expression levels also showed strong reproducibility (R^2^ = 0.79 - 0.97) (Fig. 3*C*, S2*A-D*). We also compared expression profiles across conditions using technical replicates of individual serum samples, showing that 92-98% of genes were significantly correlated between conditions (FDR < 0.1; Fig. S*2B-D*).

In these experiments, we assessed both gene-level variability (differential expression) and sample-level variability (rank consistency), with the former being a more stringent metric that is better suited for assessing signal to noise for subtle biomarker signatues.^39^ These data showed that the LC-iCAP has strong analytical reproducibility and can detect signal above background across various conditions used in assay development. Therefore, the variability seen in the earlier proof-of-concept experiment (Fig. 2B) likely arises from pre-analytical sources, not analytical instability.

#### Validation of Hypoxia Signaling as a Generalizable LC-iCAP readout

To evaluate the generalizability of the hypoxia response identified in Cohort 1, we analyzed the LC-iCAP readout using the pooled serum standards, composed of 16 individual samples, 12 of which were distinct from those in the original Cohort 1 (Fig. 2*B*, bottom). First, RNA-seq data from two pooled serum control experiments (n=8; Datafile 2) were combined and analyzed for differential expression between case versus control groups. GSEA identified hypoxia as the most enriched pathway (adjusted p-value < 0.05, Fig. S4). Next, we repeated the LC-iCAP with the pooled serum standards using a different normal bronchial epithelial cell line (NuLi1) as indicator cells. We identified 61 genes that were significantly differentially expressed in both LC-iCAP RNA-seq experiments (using 16HBE and NuLi1 cells), with expression changes showing strong correlation between cell types (R 0.77, Fig. 4*A*, *left*). STRING network analysis of the 47 coherently up-regulated genes revealed high network connectivity and enrichment of HIF1A/response to hypoxia and other processes (FDR < 0.001, Fig. 4*A*, *right*).

**Figure 4.**
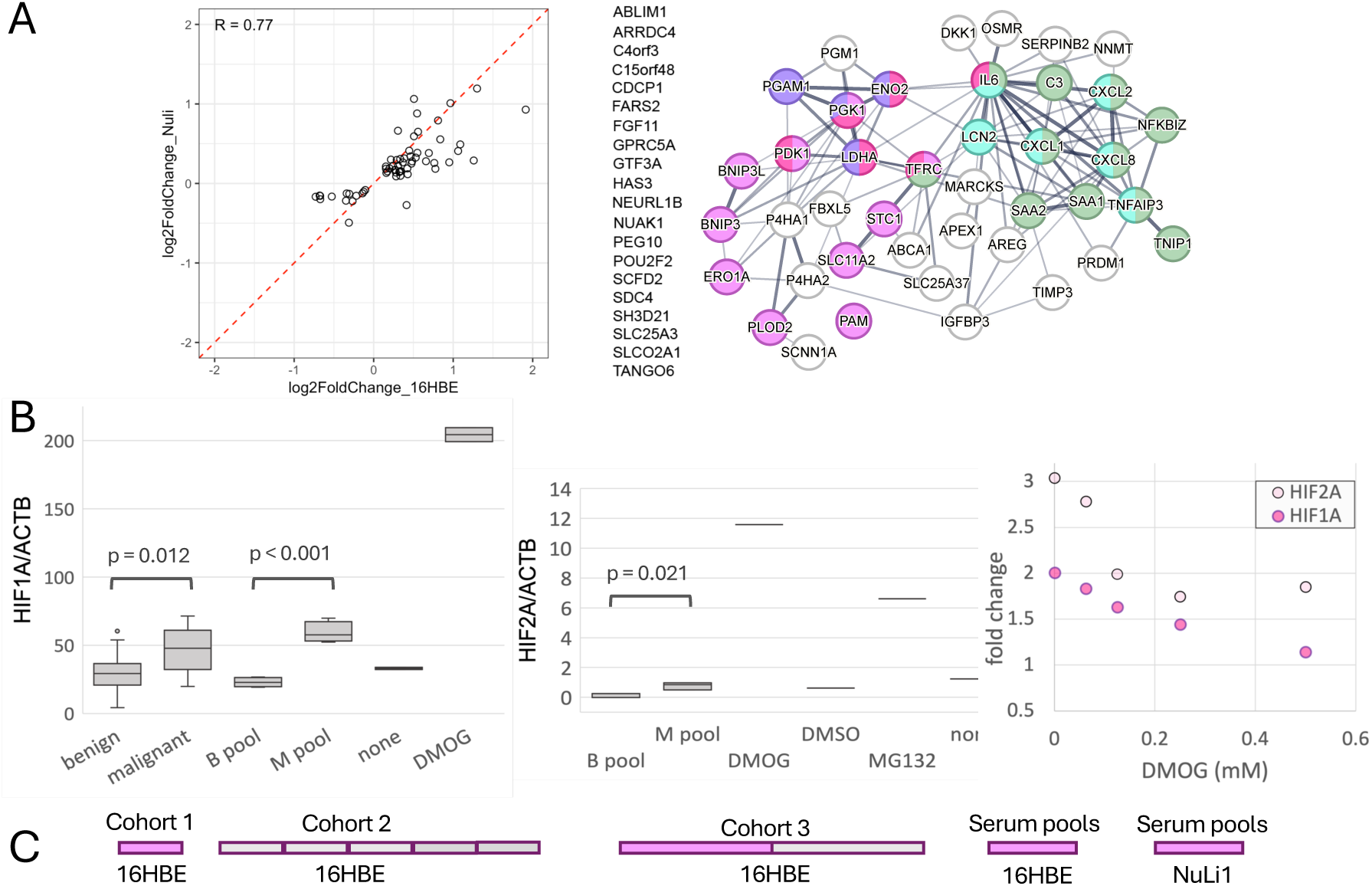
The LC-iCAP response to case versus control serum is enriched for HIF1A-mediated hypoxia signaling in multiple bronchial epithelial cell lines. ***A.*** *Left,* comparison of case versus control differential expression in LC-iCAP RNAseq data using two different bronchial epithelial indicator cell lines (16HBE and NuLi-1), each with 4 replicates of pooled serum controls. The 61 genes with differential expression in both cell types are shown (FDR < 0.1). *Right,* STRING network analysis of the upregulated genes show enrichment of network connectivity (p-value <1E-16) and enrichment of annotations including HIF1A/response to hypoxia (*red/pink*), glycolysis (*purple*), and IL-17 signaling/inflammation (*cyan/green*) (FDR < 0.001). Edge thickness represent strength of interactions. Genes with no connections or annotations are listed at the *left*. ***B.*** Results of western blot analysis showing upregulation of HIF1A and HIF2A transcription factors in the LC-iCAP with 16HBE cells in response to case versus control serum. *Left and middle,* box plots showing levels of HIF1A or HIF2A normalized to actin across 4 replicates of pooled serum controls (M pool and B pool) and across 28 different individual serum samples including 14 of each class (malignant and benign). The 28 Individual samples included the 12 samples used to make the pools. Positive controls were one or two replicates each of DMOG or Mg132 versus DMSO or no stimulus. *Right*, one replicate of each serum pool was analyzed in the LC-iCAP with increasing concentrations of DMOG showing that DMOG dampens case versus control differential expression for both factors. Western blot images are in Figure S5. ***C.*** Case versus control differential expression and enrichment of ‘HIF1A signaling’ and ‘response to hypoxia’ were observed in various LC-iCAP RNAseq experimental batches in this study *(FDR < 0.01*), whereas others had very weak or no detectable differential expression (*gray*). All LC-iCAP models with significant performance in this study used features selected from one or both of the hypoxia-enriched batches of cohort 1 or 3.

Towards elucidating the response mechanism, we compared levels of hypoxia-responsive transcription factors HIF1A and HIF2A between case and control classes in the LC-iCAP using quantitative western blotting on both serum pools and individual samples. Significantly higher levels of both factors were observed with case versus control sera, an effect reduced by the addition of DMOG, a known HIF1A stabilizer (Fig. 4*B*). Collectively, these results support that hypoxia signaling is a generalizable marker of lung cancer in the LC-iCAP and suggest the involvement of HIF1A and HIF2A in the response observed in the indicator cells.

Throughout assay development, several LC-iCAP experimental batches had enrichment of ‘HIF1A signaling’ and ‘response to hypoxia’ GO molecular processes in case versus control differentially expressed genes (Fig. 4*C*, Data file 2), whereas batches without enrichment had very few if any differentially expressed genes with FDR < 0.1.

#### Selection of model features for assay validation

Although LC-iCAP RNA-seq data is well-suited for assay optimization and development, it is not sufficiently high throughput (HTP) for assay deployment. Therefore, we used the LC-iCAP RNA-seq data to select 85 features for a final model training and validation study with LC-iCAP NanoString Plexset data to transition to a HTP readout. This cross-platform approach reduces the risk of model overfitting due to any platform-specific biases and increases the robustness of the final model.

To do this, we conducted a large-scale model parameterization study with LC-iCAP RNA-seq data and then selected features from 3 top models with optimal parameters (outlined in Fig. 5). We used data from cohorts 1-3 (Table I), filtered samples based on sample and data quality (see Methods) and partitioned them into a training and held-out validation set. Next, we trained 52 RF models with 20 seeds each, tested them on the validation set and ranked model parameters based on performance. Parameters explored were evidence-based: 1) Feature selection was based on case versus control differential expression either across all samples or within sample subsets, an approach taken to improve signal detection in the presence pre-analytical variability in the data, and 2) Optional sample filters to remove either non-smokers or patients with low lung function were applied based on our findings (Fig. S6) or those of other studies.^40^ Notably, the highest-performing models consistently applied one or both sample filters and favored features identified from sample subsets with strong differential expression rather than those derived from the full training set, suggesting the importance of accounting for clinical heterogeneity in model development.

**Figure 5.**
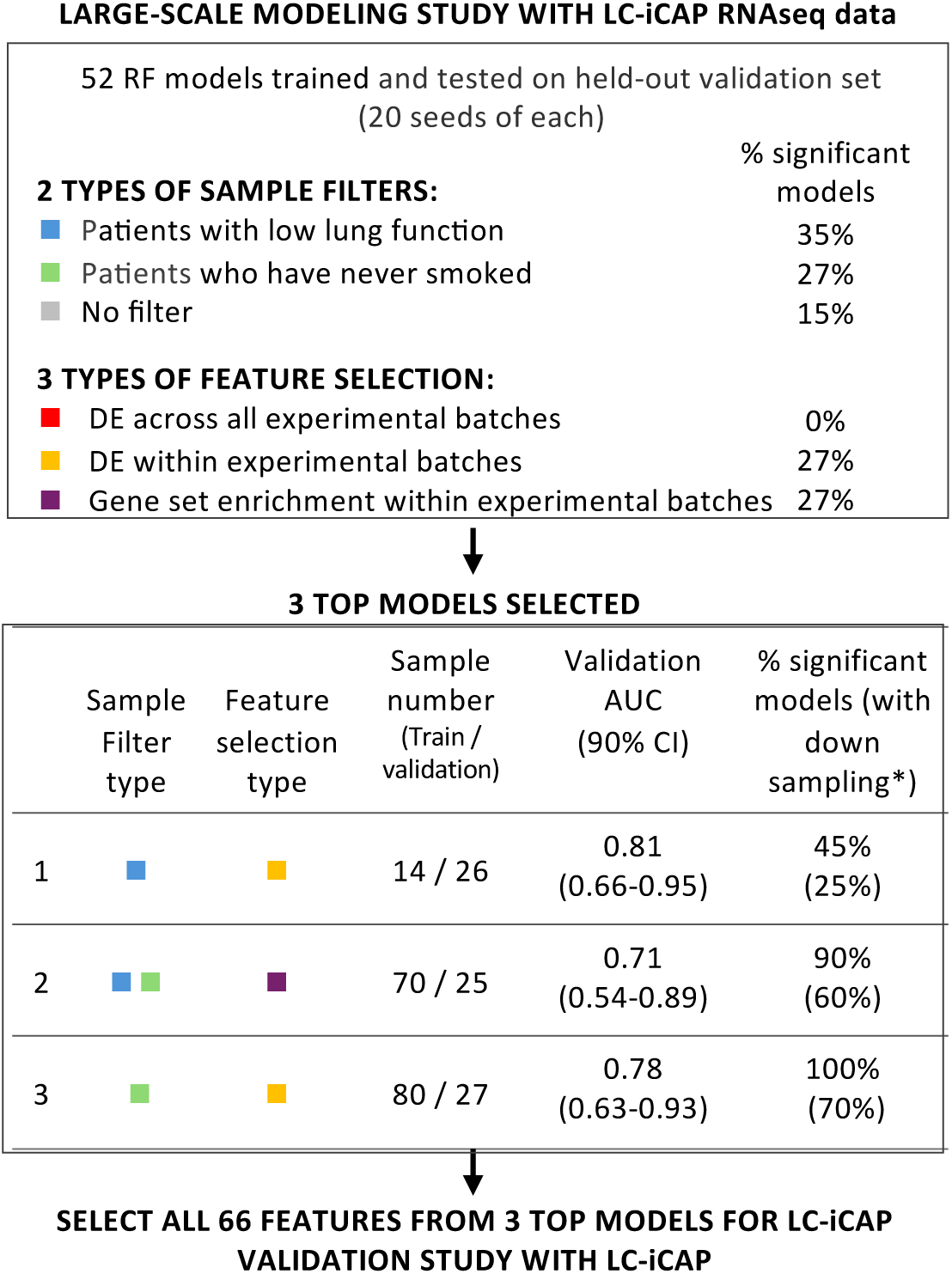
Summary of a large-scale modeling study with LC-iCAP RNA-seq data to identify a subset of optimal features for final model development with LC-iCAP NanoString Plexset data. First, 52 RF models were trained using various parameters, each with 20 seeds, and tested on a held-out validation set. Results are summarized (*top*). Next, three top performing models were chosen (*bottom*) and the 66 features from these models were selected for the validation study. % significant models is the percentage of models in the category having at least one seed with significant performance. *model training was performed with and without down sampling to balance classes within each experimental batch. *DE*, differential expression.

Three top models were selected with high performance, including one with AUC of 0.78 (90% CI 0.63-0.93) with 100% sensitivity and 60% specificity (Fig. 5 bottom). A list of 85 features was generated for the LC-iCAP validation study composed of 66 features selected from multiple seeds of the 3 top models and an additional 19 genes from the proof-of-concept pilot model not yet in the list (Data file 3).

### LC-iCAP Validation

In this stage, we generated LC-iCAP NanoString Plexset data for the 85 candidate features selected in the previous stage and used these data for final LC-iCAP model development and blind testing. The transition from RNA-seq to Nanostring Plexset, a high-throughput platform for analysis of up to 96 genes across 96 samples per batch, was made not only to minimize platform-specific biases in the model, but to initiate the development of a high-throughput assay configuration suitable for clinical deployment.

#### Generating LC-iCAP data with high-throughput readout

We generated LC-iCAP data using the NanoString PlexSet platform for model training across cohorts 1–3 (Table I, Data File 4). After merging and normalizing the data, we applied filtering based on sample and technical quality, and patient characteristics, resulting in a final dataset of 97 samples for modeling (Fig. 6). This included: 1) Excluding data from the small number of ‘never smokers’ based on the results of assay optimization, which also improved the relevance of this study to lung cancer screening which focuses on current or former smokers only^7^, 2) Removing data from samples with hemolysis using an established threshold,^17^ which was implemented in assay optimization, and 3) Excluding low-quality samples with storage times > 10 years based on our analysis showing an effect of storage times > 10 years on gene expression and HIF1A activity in the LC-iCAP (Fig. S7, S8). Our data are consistent with studies showing that biomarkers degrade with storage, but that most proteins and metabolites tested are stable for up to 7.3 years.^41,42^

**Figure 6.**
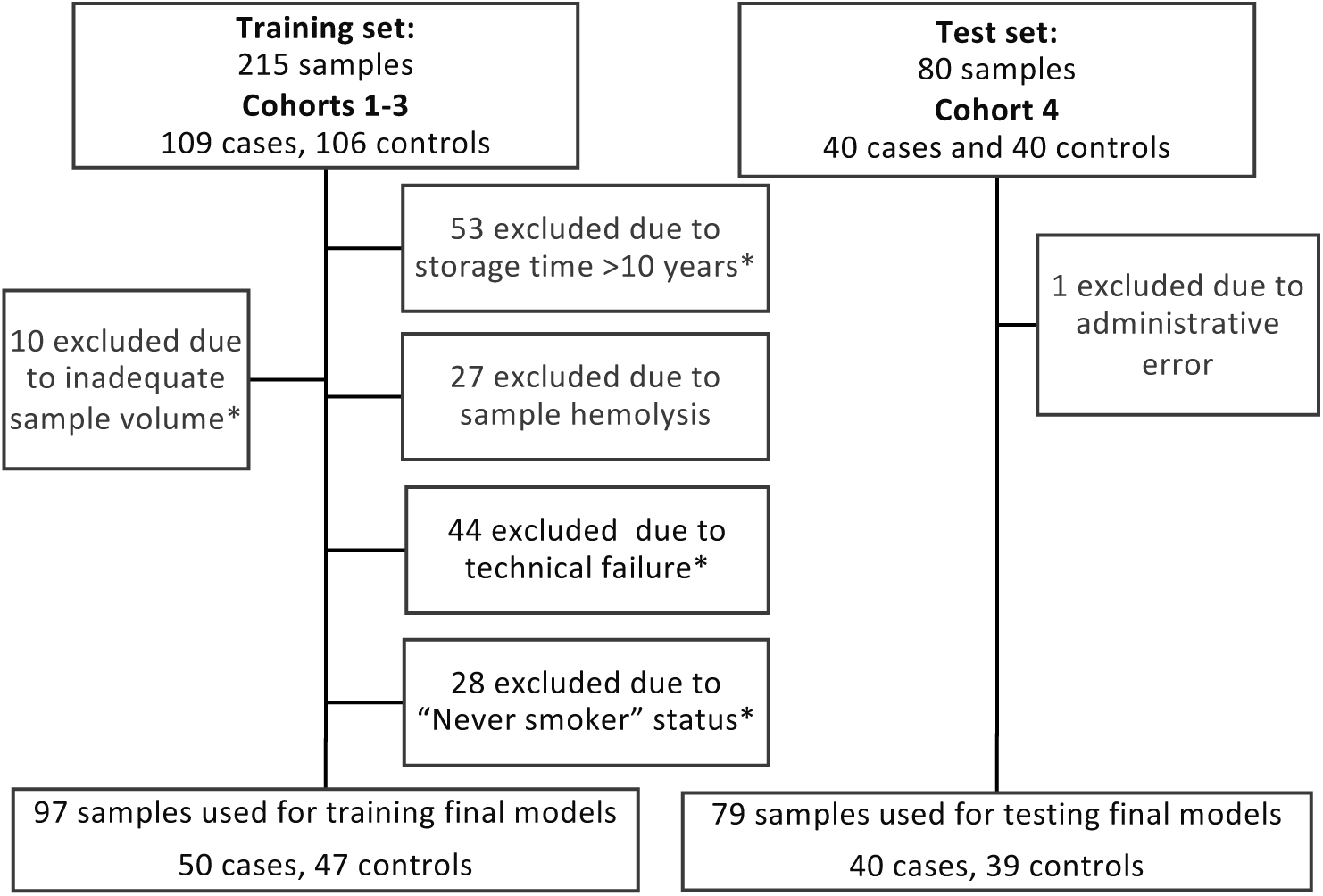
Retrospective sample flow diagram for LC-iCAP validation study. Cohorts 1-3 were used for model training and cohort 4 was used as a blind test set. The training set and blind test set had temporal independence; test samples were collected 9 years later and assayed 1 year later than the training samples. Sample numbers are not cumulative as samples can belong to multiple exclusion groups. *Samples indicated were used for LC-iCAP development and/or feature selection.

After data generation, we tested LC-iCAP Plexset reproducibility by hierarchical clustering using the standard control data and found reproducibility of expression and differential expression across the 5 Plexset plates (Fig. S9, code set lot 1 samples).

#### Final LC-iCAP model training

We used data for the 97 samples as a training set for model development. Selection of parameters and hyperparameters and estimation of model performances was done with nested cross-validation, an approach that has been demonstrated to generate unbiased performance estimates with small sample sizes.^35^ Most parameters tested were based on the results of assay optimization in Figure 5, except here, patient smoking status (current or former) was included as a covariate in the models.

#### Final blind testing of LC-iCAP model

For blind testing of locked-down models, a new set of 80-samples from Vanderbilt University was added to the study (Cohort 4, Tables I and II). The patient characteristics matched those of the training set, except all IPNs had pretest risk of malignancy between 5-65% calculated using the Mayo Clinic model, and all had high serum quality with storage times between 1-4 years. Additionally, samples of the blind set had temporal independence from the training set, with a collection window that was 9 years later and LC-iCAP processing that was 1 year later, adding more rigor to the validation than concurrent collection and processing.^43^

The blind samples were analyzed by LC-iCAP-NanoString Plexset and data quality was checked by analysis of 31 DMOG chemical controls run with code set lot 1(training) and code set lot 2 and (blind test set samples). This identified a miscalibration between the training set and test sets that measurably affected 50 of the 85 genes (Fig. S9, Data file 4), which was attributed to technical failure of normalization between the code set manufacture batches. To overcome this, we trained models after omission of the 50 mis-calibrated genes and four models were tested on the blind set using either RF or GLM. Only the RF models (M3 and M4) had significant performance (Fig. S10, Fig. 7*B*, Fig. S11*B*) showing better robustness of the RF ensemble model in preserving predictive signal from the remaining genes despite the calibration issue. For comparison, a total of five models were tested prior to gene exclusion, none of which showed no significant performance on the blind set (data not shown). Note that during testing of the 9 models, test samples were kept blind to data scientist and parameter selection was based only on mitigating miscalibration.

**Figure 7.**
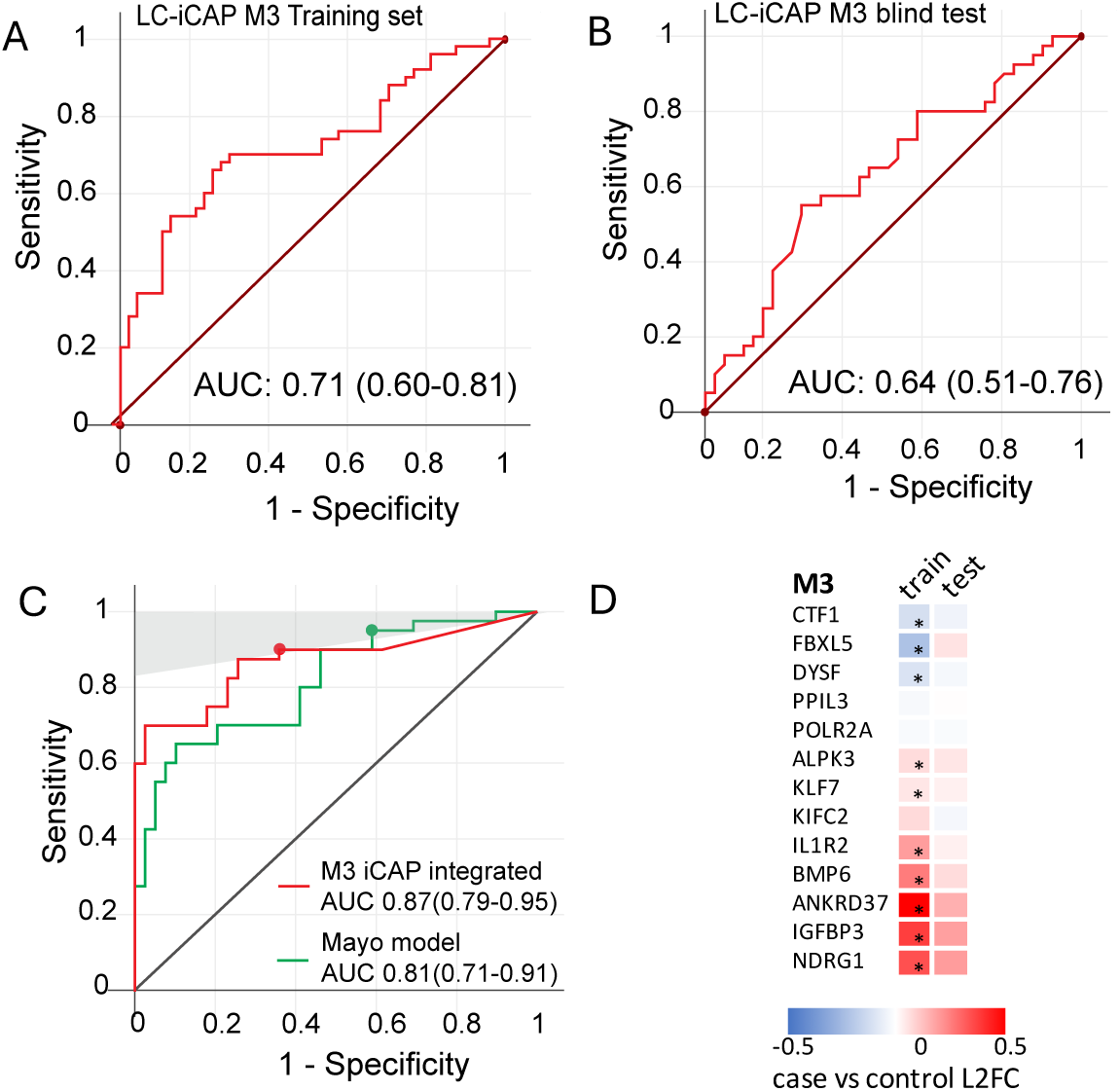
ROC curves showing performance of the M3 LC-iCAP model and the M3 iCAP-integrated classifier in temporal blind validation on the test set. ***A and B*** The M3 LC-iCAP model demonstrated significant performance based on out-of-bag (OOB) predictions from the training set (A) and on an independent blind test set (B) ***C*.** The iCAP-integrated classifier outperforms the Mayo Clinic model on the test set. The *shaded* region corresponds to ≥ 95% NPV using a 25% prevalence estimated in the clinical population. The model performances were compared at cut points with clinical utility as rule-out tests (with ≥ 95% NPV and ≥ 90% sensitivity) (*colored nodes*) (corresponding to 96.1% NPV, 41% specificity and 95% sensitivity for the Mayo model and 95.1% NPV, 64% specificity and 90% sensitivity for the iCAP integrated classifier). The integrated classifier had significantly higher specificity than the Mayo model (exact binomial test p-value = 0.049) at the cut point. AUCs are shown with 95% confidence intervals in brackets. ***D.*** Heatmap of LC-iCAP NanoString Plexset data showing case versus control differential expression for genes of model M3. For samples of the training and test sets, median differential expression values are shown (except for one gene below limit of detection in >50% of samples). Genes with significant differential expression are indicated with asterisks (Mann Whitney U test p-values <0.05). Training versus test set differential expression profiles had a strong Pearson correlation coefficient of 0.8 (*p*-value 0.001) showing good reproducibility of expression between training and blind test sets. L2FC, log2 fold change.

After blind testing of the two RF models, gene features of both models were analyzed for case versus control differential expression in the training and blind test sets. Comparison of training and test set profiles showed significant correlation for both M3 (R = 0.8, Fig 7*D*) and M4 (R = 0.73 Fig. S11*D*). This supports the biological validity and generalizability of the gene signatures used by the models, reinforcing confidence that the models are not simply overfitting the training data. However, no genes had significant differential expression in both sample sets suggesting patient-to-patient variation and the importance of using a multivariate signature.

#### Exploring clinical implementation of the LC-iCAP

To explore the potential clinical utility of the LC-iCAP, a prototype clinical version of the assay was developed after blind testing by integrating the final LC-iCAP model with the Mayo Clinic model^33^ using an approach similar to that used for the Nodify XL2 test offered by Biodesix^10^ as described in Figure S12. Performance of this ‘iCAP integrated classifier’ was compared to that of the Mayo Clinic model at specific cut points on the ROC curves with clinical utility to rule-out cancer (corresponding to sensitivity ≥ 90% and NPV ≥ 95% using an estimated cancer prevalence of 25% in a community pulmonary practice).^2,3^ At this threshold, the specificity of the iCAP integrated classifier was significantly better than that of the Mayo Clinic model suggesting clinical utility of the LC-iCAP (McNemar exact binomial test p-value^34^ = 0.049) (Fig. 7*C*). Similar results were observed with the other validated LC-iCAP model (Fig. S11*C*). Because the models were compared at ROC curve points with specific performance metrics (rather than absolute probability estimates), model calibration was not required and when implemented had no effect on the results (data not shown). The parameters of the integrated classifier were selected using the blind samples; while the study demonstrates the potential performance of the LC-iCAP model, the fixed constants of the integrated classifier remain to be independently validated.

## Discussion

Blood biomarkers are needed for the early detection of diseases to improve outcomes, but their very low abundance and high levels of noise present significant technical challenges. We are developing the iCAP, a biosensor assay for blood-based diagnostics to overcome this issue by capitalizing on the evolved ability of cells to detect and respond to weak signals in noisy environments. The assay works by using cultured cells to detect disease-related molecules in blood and analyzing the gene expression response using machine learning tools for disease classification. Here, we developed the LC-iCAP, a multivariate blood test for patients with IPNs identified by CT scans to improve malignancy risk assessment and help those with benign nodules avoid invasive biopsies while directing further diagnostic efforts towards those with lung cancer.

The iCAP transforms disease signals in blood into a standardized gene expression readout from cells, enabling applying well-established tools from cell biology to the emerging field of blood-based diagnostics. This could accelerate the field much like how cell-based assays advanced drug development.^44–46^

Here, the cell-based approach enabled statistical enrichment analysis of the case versus control differential readout, which identified a lung cancer-specific ‘response to hypoxia’. It also enabled biochemical analyses, which implicated transcription factors HIF1A and HIF2A in mediating the response (Fig. 4). Consistently, all LC-iCAP classifiers with significant performance used hypoxia-related gene features that were selected from specific experimental batches with differential hypoxia enrichment (Fig. 4*C*). These finding align with hypoxia as a well-documented aspect of lung cancer and other malignancies,^38,47,48^ and suggests that the LC-iCAP readout reflects blood analytes associated with the tumor status.

The cell-based approach also enabled us to develop standardized biological and chemical controls with quantitative multicomponent readouts that we used to optimize assay and model parameters and detect and control for sources of unwanted variation in assay development. The controls were also used to monitor assay performance, identifying two technical failures during development (Fig. S3 and S9), highlighting the robustness of the standards as built-in fail safes. The standards were also used for analytical validation, showing reproducibility of case versus control differential expression in LC-iCAP across various conditions (Fig. S2, 3*C,* and S9). This included stability of the 16HBE indicator cells across cell lineages, which is supported by their use in various studies over the last 35 years,^49^ and the generalizability of the differential hypoxia response across two distinct bronchial epithelial indicator cell lines with different origins (16HBE and NuLi1). These results indicate that the cell-based assay readout possess the stability and robustness required for clinical deployment. These standard controls are valuable tools for monitoring the cell readout during assay deployment to detect and mitigate any assay failures or drift.

Despite good analytical reproducibility, we identified noise in the assay readout as the differential response to hypoxia underlying disease classification was not observed across all sample subsets, and effect sizes were small and variable (Fig. 4C, 7*D* and S11*D*). We demonstrated that this noise stemmed from patient heterogeneity (e.g., variability in lung function) and differences in serum quality (e.g., storage duration) (Fig. S6–S8). These findings motivated selective sample exclusions to improve data quality, as well as the use of a clinical covariate in modeling to enhance assay performance. It also motivated our use of RF modeling, ensemble approach enabling detection of multiple disease signatures. These data warn of the potential impact of variable sample storage times in other retrospective studies. They also highlight the value of multivariate ensemble models in diagnostics for robust and generalizable outcomes, and for next-generation applications in disease stratification.

To develop the LC-iCAP for lung nodule management, we first constrained gene expression features from ∼20,000 down to 85 using a modeling-based approach with LC-iCAP RNA-seq data.

The 85 genes were the features from 4 models including a pilot model that was trained on 12 samples and had significant performance when tested on a held-out validation set of 103 samples (AUC 0.62) (Fig. 2*A*), and 3 optimized models with improved performance on a held-out set (AUCs of 0.71-0.91) (Fig. 5). The 85 gene features and a smoking status covariate were then used to make a high-throughput version of the assay compatible with clinical deployment using samples from patients with IPNs who were either current or former smokers (Table II). The models were trained by nested cross-validation and tested by blind validation. The test set was temporally independent from the training set in both sample collection and processing, enhancing the rigor of the validation compared to internal validation where one dataset is randomly split into a training and test set.^43^ Two final models had significant performance on a blind set with AUCs of 0.64 (95% CI: 0.51–0.76) and 0.62 (95% CI: 0.50-0.75) (Fig. 7*C* and S11*C*). The generation of significant models each using overlapping subsets of genes, reflects the coregulatory nature of cellular gene expression networks and highlights the potential value of using a cell-based response in diagnostics to capture patient diversity.

**Table II.**
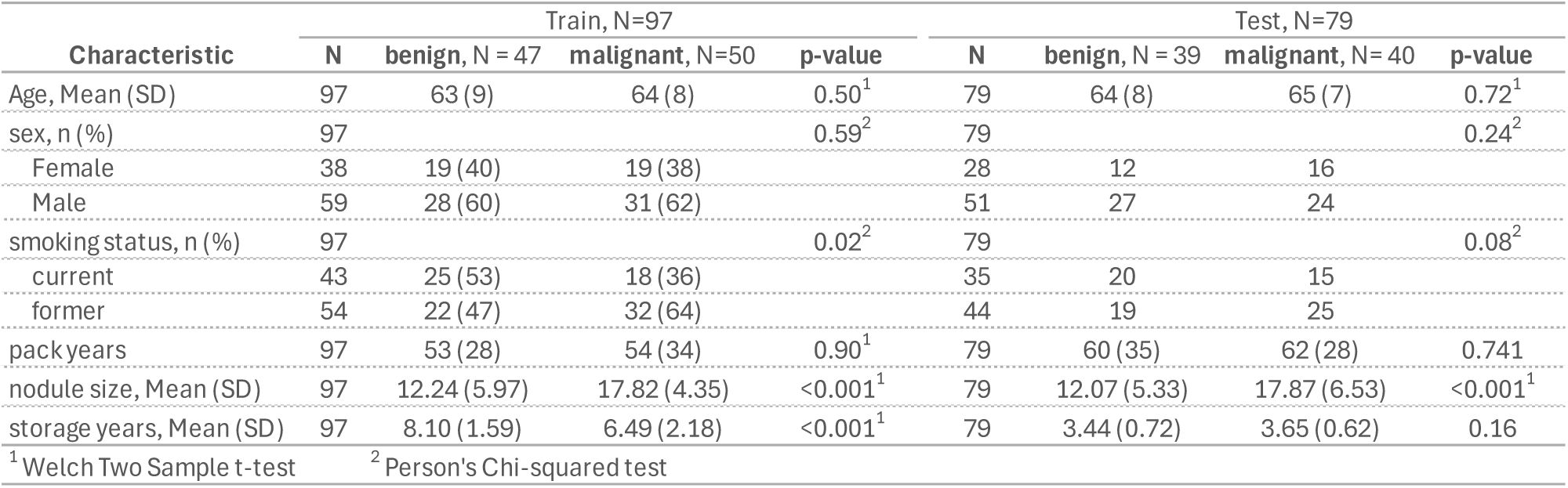
Participant demographics for LC-iCAP validation study.

After blind testing we assessed the potential clinical utility of the final LC-iCAP model by integrating it with the Mayo clinic model using a method previously developed for deploying the Nodify XL2 test by Biodesix^10^ (Fig. S12). The ‘iCAP integrated classifier’ performance was compared to that of the Mayo model at specific cut points yielding ≥ 95% NPV using a 25% prevalence in the intended use population.^2,3^ This threshold was selected to maximize clinical utility as a rule-out test to discriminate nodules that have ≤ 5% risk of cancer that can be diverted to surveillance from those with higher risk that require further testing. At this threshold, the LC-iCAP integrated classifier had 64% specificity and 90% sensitivity and significantly better performance than the Mayo model suggesting clinical utility (Fig. 7*C*).

In a clinical setting where a rule-out test is used to guide patient care, specificity at the cut point indicates the percentage of patients with benign nodules who would be directed to surveillance, thus avoiding potentially invasive follow-up procedures. The specificity of the LC-iCAP integrated classifier at 64% was 1.45-1.9X better than that reported for the two other CT-integrated blood tests from Biodesix and Magarray at similar NPV thresholds, (44% and 33%, respectively). ^9,10^ This suggests actionable results for a greater number of patients with benign nodules using the LC-iCAP. Sensitivity, reflecting the percentage of patients with malignant nodules who would be correctly directed to follow-up testing, was 90% for the iCAP integrated classifier. Although this is lower than that of the other tests (97% and 94%), the iCAP integrated classifier would have a 95% NPV in the intended use population, attributable to its higher specificity at the cut point, thus not substantially increasing harm.

While the data are promising, this study has limitations. Our data suggest that the use of archived samples with variable storage times may have negatively affected model performance (Fig. S7, S8). In addition, to overcome a NanoString Plexset technical failure, a total of 9 models were tested on the blind test set (5 before and 4 after omission of affected genes) and two had significant performance. However, because all models were constrained to the same 85 pre-defined features, the risk of spurious findings due to multiple hypothesis testing is greatly reduced compared to selection from the entire transcriptome. Finally, although temporal validation is rigorous, external validation should be performed. Our next step is to conduct a multi-site prospective study with a larger sample size to further improve model performance and robustness, followed by a clinical utility study as recommended by the American Thoracic Society.^8^

The iCAP is a multivalent platform with a multivariate readout. Requiring only a small amount of serum, all analytical steps are scalable to 96-well format in assay deployment. Therefore, the iCAP could have utility as a next-generation platform for cancer screening including multi-cancer early detection (MCED). This could involve either using an array of indicator cells or tuning the LC-iCAP to detect multiple cancer types. Supporting this, HIF1A-mediated hypoxia response in the LC-iCAP readout has central roles in general tumor biology.^47,48^ Notably, hypoxic tumors are more likely to metastasize and are less likely to respond to treatment^48^ and to our knowledge, blood biomarkers of tumor hypoxia have not yet been identified. Future studies include exploring the use of fluorescent reporters and single cell analysis to further simplify and amplify the LC-iCAP readout, as well as scaling all analytical steps to 96-well configuration using tools already in use for diagnostics such as QuantiGene Plex® instead of NanoString Plexset.

To achieve the Cancer Moonshot initiative’s goal of accurate early detection with minimal overdiagnosis and missed cases, relying on only one test is unlikely to yield optimal clinical utility. Multimodal approaches, which combine multiple orthogonal tests each with independent performance, can outperform individual tests in isolation.^34,50^ The iCAP is a non-conventional approach that is complementary to other more traditional tests and thus is well-suited for combinatorial diagnostics. Through collaborative efforts, just as the power of combining treatments has revolutionized therapeutics, the integration of diverse diagnostic modalities holds the promise of transforming diagnostics as well.

## Data Availability

All data produced in the present work are either contained in the manuscript or supplementary tables and figures except for complete Raw and processed RNAseq data. Raw and processed RNAseq data have been submitted to GEO repository and will be publicly available after peer review.

## Funding and Acknowledgements

Research reported in this publication was supported by the National Cancer Institute (NCI) and National Institute of Aging (NIA) of the National Institutes of Health (NIH) under Award Numbers R43CA203455, R44CA203455 and R44AG051282 to PreCyte, the National Institute of Environmental Health Sciences (NIEHS) of the NIH under Award Number P30ES013508 to the University of Pennsylvania, and the NCI of the NIH under award number P30CA068485 and NCI-U01CA152662 to Vanderbilt University. The content is solely the responsibility of the authors and does not necessarily represent the official views of the NIH.

## Author contribution

JDA, JJS, and RJL and conceived the study, JDB and JJS designed the experiments. AV, PM, and SD selected and provided samples and insights related to clinical need. JDB, LRM and YQ conducted the experimental work and collected the data. Data analysis was performed by FJD, GAW, JDB, JJS, MDD, SAD, SD and YQ. The manuscript was written by JJS. All authors reviewed and approved the final manuscript. JDA, JDB, JJS and RJL supervised the project and provided critical insights throughout the study.

