## Supplementary Figures for "A multivariate cell-based assay for blood-based diagnostics enhances lung cancer risk stratification"

### Figure S1. LC- iCAP optimization with case and control serum pools

LC-iCAP optimization studies were conducted by comparing the assay readout with different experimental parameters to identify the optimal conditions. The optimization studies described here used technical replicates of pooled patient serum samples. For each experiment, unless otherwise stated, four technical replicates of case and control pools were analyzed in the iCAP under each condition and gene expression readout was measured by NanoString analysis (using the discovery gene set of 88 candidate genes or by RNAseq analysis (described in the Methods Section). Using these data, malignant versus benign class separation was compared across conditions using: 1) Principal component analysis (PCA), a method to summarize collective activity of multiple genes across different samples, 2) Plots comparing magnitude of log2 fold change (L2FC) of differentially expressed genes between pairs of conditions, and 3) Volcano plots that show magnitude and significance of differentially expressed genes (DEGs) under one condition.

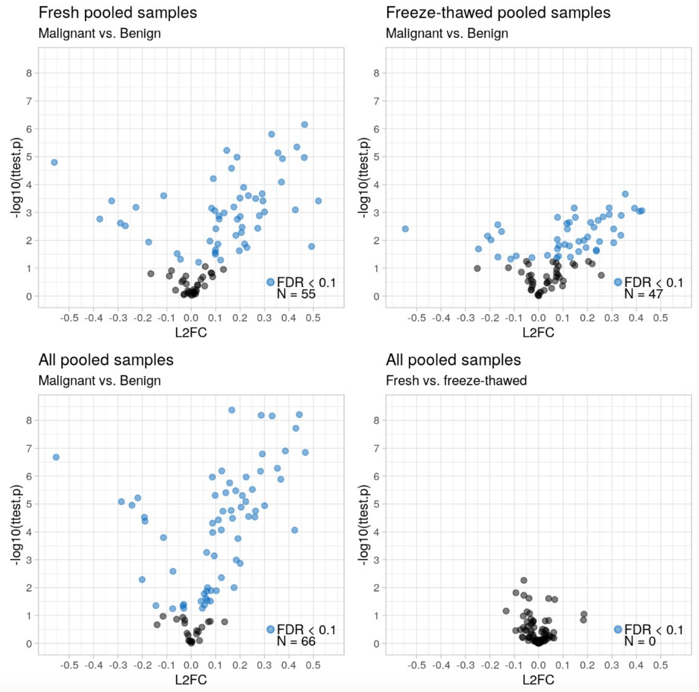


**Fig. S1*A*. The LC-iCAP with once- and twice-thawed serum pools.** *Top,* volcano plots of LC-iCAP Nanostring data with once-thawed (fresh) versus twice-thawed serum pools (freeze-thawed) show that the number of significantly differentially expressed genes is similar under both conditions, but a second thaw appears to lower significance of differential expression. *Bottom*, combining the data from both experiments yields lower p-values (*left*), and comparison of once- versus twice-thawed serum shows no significant differential expression (*right*). These data show that once- and twice-thawed serum yield similar number of genes with significant differential expression in the LC-iCAP but that data with once-thawed serum has a stronger confidence. These data support the use of twice thawed serum for optimization studies described below.

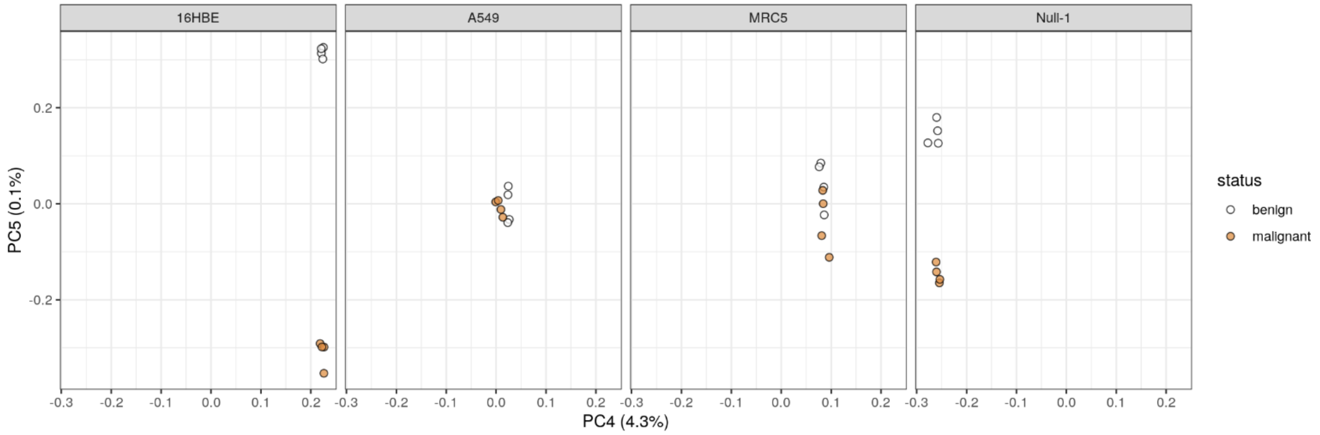


**Fig. S1*B.* LC-iCAP indicator cell type optimization**. PCA analysis of LC-iCAP Nanostring data comparing class separation with 4 different indicator cell types. A549 (derived from malignant lung tissue) and MRC5 fibroblast cell lines show no class separation, whereas Nuli-1 and 16HBE had noticeable class separation.

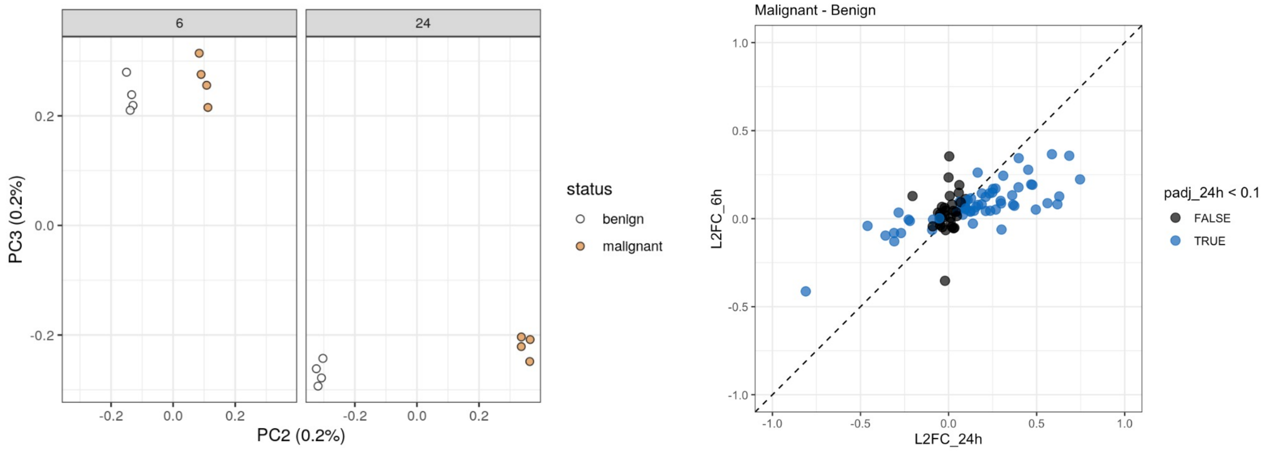


**Fig. S1*C.* LC-iCAP serum incubation time optimization.** ***Left,*** PCA analysis of LC-iCAP Nanostring data comparing class separation with 6 h versus 24 h serum incubation times. Class separation was observed under both conditions, but effect was greater after 24 h. ***Right,*** Comparison of magnitude of differential gene expression between 6 h and 24 h incubation shows that direction of change is coherent across the two incubation times for most genes, but magnitude is higher for 24 h condition. 37 and 57 genes of 88 genes on the panel are significantly DEG for 6 h and 24 h, respectively with adjusted p-values <0.1). These data show that 24 h incubation is optimal, and that assay readout is robust to variation in incubation time.

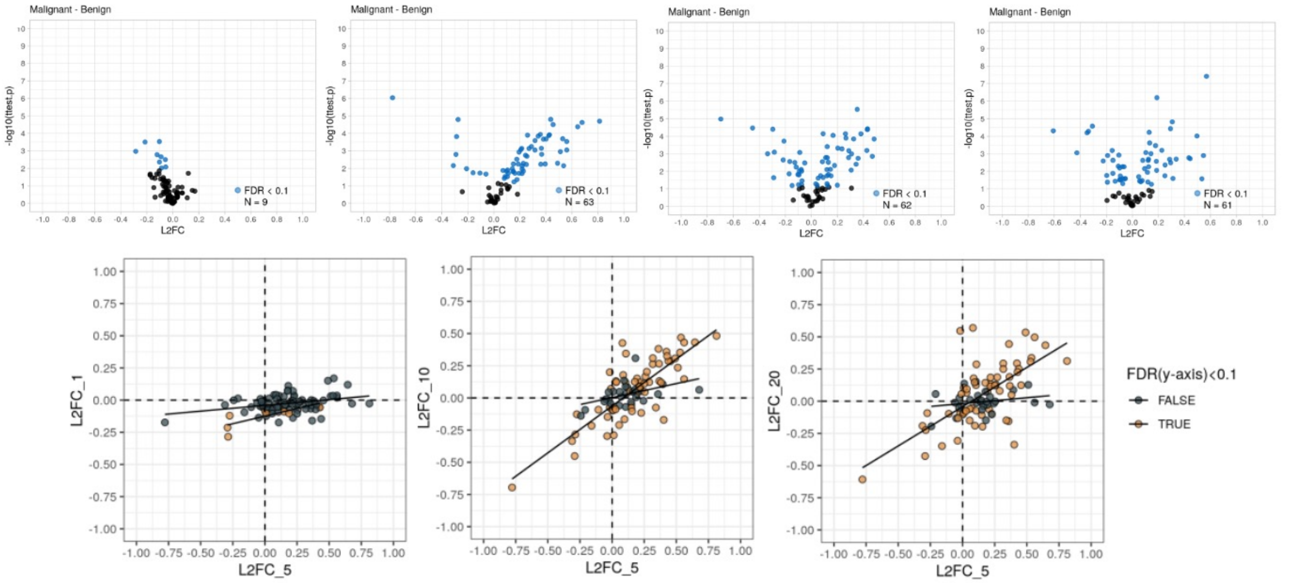


**Fig. S1*D.* LC-iCAP readout with various pooled serum concentrations**. ***Upper***, volcano plots showing differential expression in LC-iCAP Nanostring data with four different pooled serum concentrations (1, 5, 10, and 20%). 5-20% serum yields highest level of differential expression. ***Lower,*** plots comparing magnitude of differential expression between pairs of serum concentrations. Genes with significant differential expression for the condition on the Y axes are colored orange (FDR < 0.1) and fitted linear regression models of significant DEGs and other genes are shown in black. Together, data show that whereas 1% serum yields minimal differential expression in the LC-iCAP, 5%, 10% and 20% serum have stronger readouts with most genes showing similar magnitude and direction of change across serum concentrations. The standard serum concentration in the LC-iCAP is 5% and these data show the assay readout is robust to higher but not necessarily lower serum concentrations.

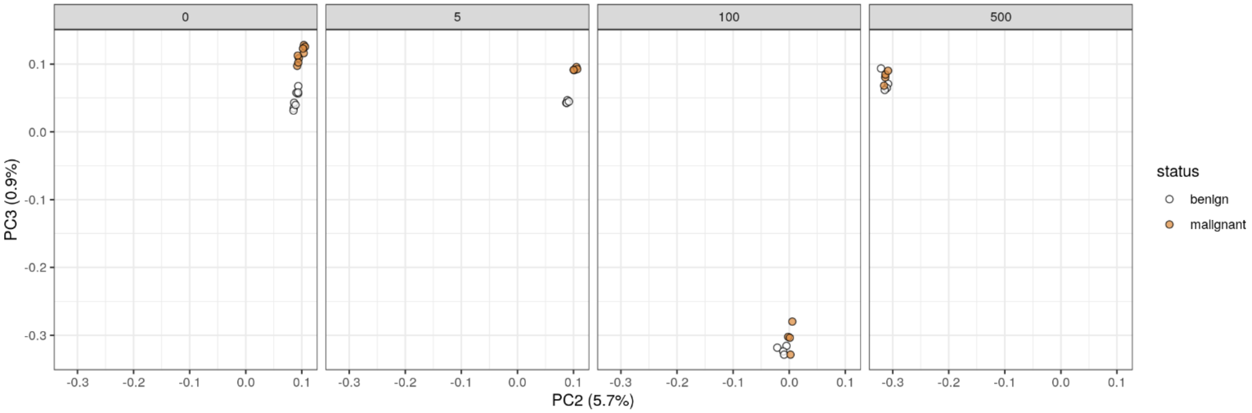


**Fig. S1*E.* Effect of trichostatin A on LC-iCAP readout.** PCA analysis of LC-iCAP Nanostring data of benign and malignant serum pools comparing class separation in the LC-iCAP with increasing concentrations of trichostatin A (TSA), an anticancer drug that stabilizes HIF1A under normoxic conditions. This experiment tests the hypothesis that TSA improves the HIF1A-mediated hypoxic response to malignant versus benign serum in the LC-iCAP. Comparison of class separation at varying concentrations of TSA (0, 5, 100, 500 nM) show no dramatic improvement of class separation in presence versus absence of the drug.

#### Supplementary Figure S2. LC iCAP Reproducibility analysis

LC-iCAP reproducibility experiments were conducted under various conditions to determine if the assay has sufficient sensitivity to detect differential expression of biomarkers in the presence of various potential sources of noise. Most experiments were performed using 4 technical replicates each of the case and control pooled patient serum standard controls per condition with a readout of differential expression measured by RNAseq or Nanostring with the development gene set. In other experiments, 12 individual serum samples including 6 each of each class were analyzed in singlet for each condition and reproducibility was measured per gene by determining correlation of gene expression profiles between the two conditions. Together these data show that LC-iCAP has sufficient sensitivity to detect differential expression in the presence of potential noise from various sources including different indicator cell lots, different LC-iCAP batches, different RNA detection platforms, and different Nanostring batches. *DEGs*, differentially expressed genes; *ECDF*, empirical cumulative distribution function; *FDR*, false discovery rate; *L2FC*, log2 fold change; *PCA*, principal component analysis; *R*, correlation coefficient.

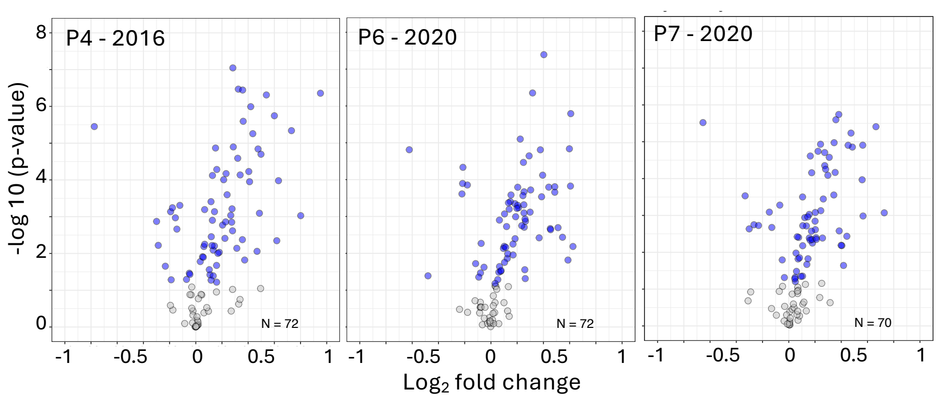


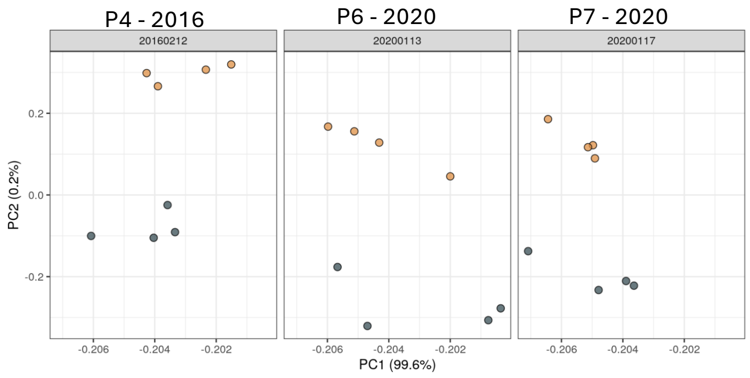


**Fig. S2*A*. LC-iCAP differential expression is reproducible across 3 different lots of indicator cell.** Comparison of LC-iCAP NanoString data for technical replicates each of case and control serum pools across three different lots of indicator cells. The three lots were from three different cell passages on three different dates spanning 4 years (See Fig. 4C for cell lineages). *Upper*, volcano plots showing similar magnitude and significance of differential expression for each passage of cells. *Lower,* PCA showing a similar level of separation of case versus control samples in the second PC (PC2) for all three cell passages (case pools are orange and control pools are gray). Additionally, we generated scatter plots with $R^2$ values showing that the three cell passages yield similar expression and differential expression variability (Fig. 4*C* of manuscript). These data show that case versus control differential expression is reproducible for 3 different indicator cell passages spanning 4 years suggesting long-term cell stability suitable for clinical assay development.

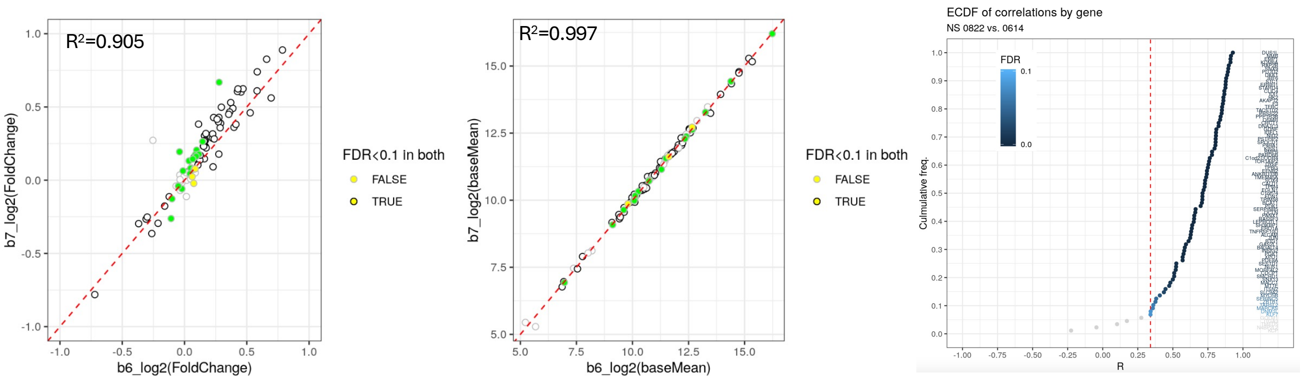


**Fig. S2*B.* LC-iCAP differential expression is reproducible across different experimental batches on different days.** Comparison of LC-iCAP Nanostring data for 4 replicates of each of case and control serum pool across two different LC-iCAP batches run on different days. *Left*, comparison of magnitude of differential expression (malignant versus benign L2FC) for each gene between the two conditions. *Middle*, comparison of average expression values for each gene between the two conditions. Genes with black edges, yellow fill and green fill are differentially expressed in both batches, batch 6 only, and batch 7 only, respectively (FDR < 0.1). $R^2$ values are shown. *Right,* LC-iCAP batch reproducibility and serum aliquot reproducibility measured with individual serum samples. LC-iCAP Nanostring data were generated with replicate aliquots of 12 serum samples (6 cases and 6 controls) on two different days. For each gene, reproducibility of expression was determined by measuring the correlation of expression profiles between the two batches. Correlation coefficients (R) were plotted on an ECDF plot showing that 94% (83 of 88) of genes had significant correlation of expression between the batches (FDR < 0.1, red dashed line). One of the 5 genes without significant correlation was below limit of detection of the NanoString assay.

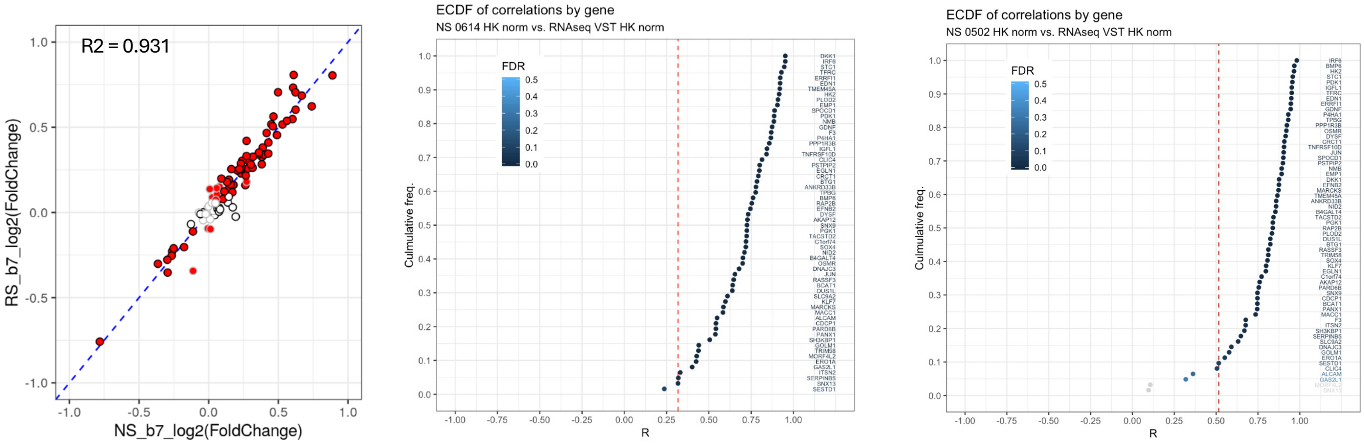


**Fig. S2*C.* LC-iCAP differential expression is reproducible across NanoString and RNAseq platforms.** Nanostring and RNAseq readouts were compared in two experiments. *Left,* four replicates each of case and control pools were analyzed by LC-iCAP with RNAseq (RS) and Nanostring (NS). For each platform, case versus control differential expression was measured for genes in the development gene set and correlations of Log2 fold changes were plotted for each gene. Significantly differentially expressed genes in RNAseq and Nanostring data are filled red and outlined in black, respectively (FDR < 0.01). *Middle and Right,* 12 individual samples (6 malignant and 6 benign) were analyzed by LC-iCAP once with RNAseq and twice with Nanostring for development genes detected by both platforms. For each gene with significant differential expression, reproducibility of expression between RNAseq and Nanostring was determined by measuring the correlation of expression profiles between the two platforms. Correlation coefficients (R) were plotted on ECDF plots showing that that 98% (61 of 62 genes) and 92% (57 of 62 genes) of genes for first and second NanoString runs had significant correlation of expression between the platforms (FDR < 0.1, red dashed line).

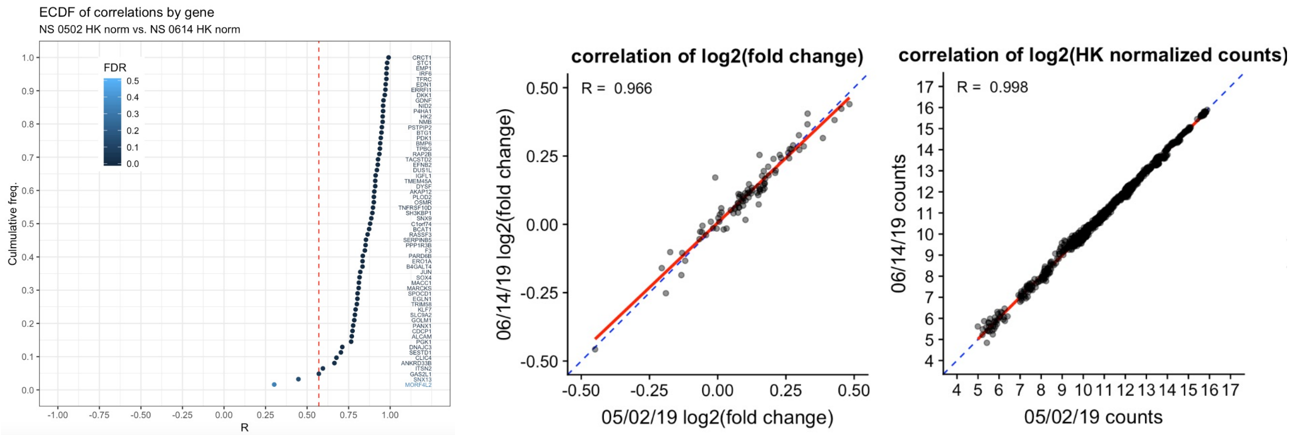


**Fig. S2*D.* LC-iCAP differential expression is reproducible across Nanostring batches.** Reproducibility between different Nanostring runs was measured by generating LC-iCAP Nanostring data for 12 individual patient samples (6 of each malignant and benign), analyzing twice in two different Nanostring batches (0502 vs 0614) and measuring correlations between the runs. *Left,* for each gene with significant differential expression in at least one run, correlation of expression patterns across the 12 samples between two Nanostring runs are plotted on an ECDF plot. 97% of genes (60 of 62 genes) were significantly correlated (FDR < 0.1, red dashed line) *Middle and right,* correlation of gene expression between the two Nanostring runs is shown using either differential expression values (log2 fold change of malignant vs benign) (*middle*) or normalized counts (*right*). Results indicating good reproducibility of gene variation between the two Nanostring batches.

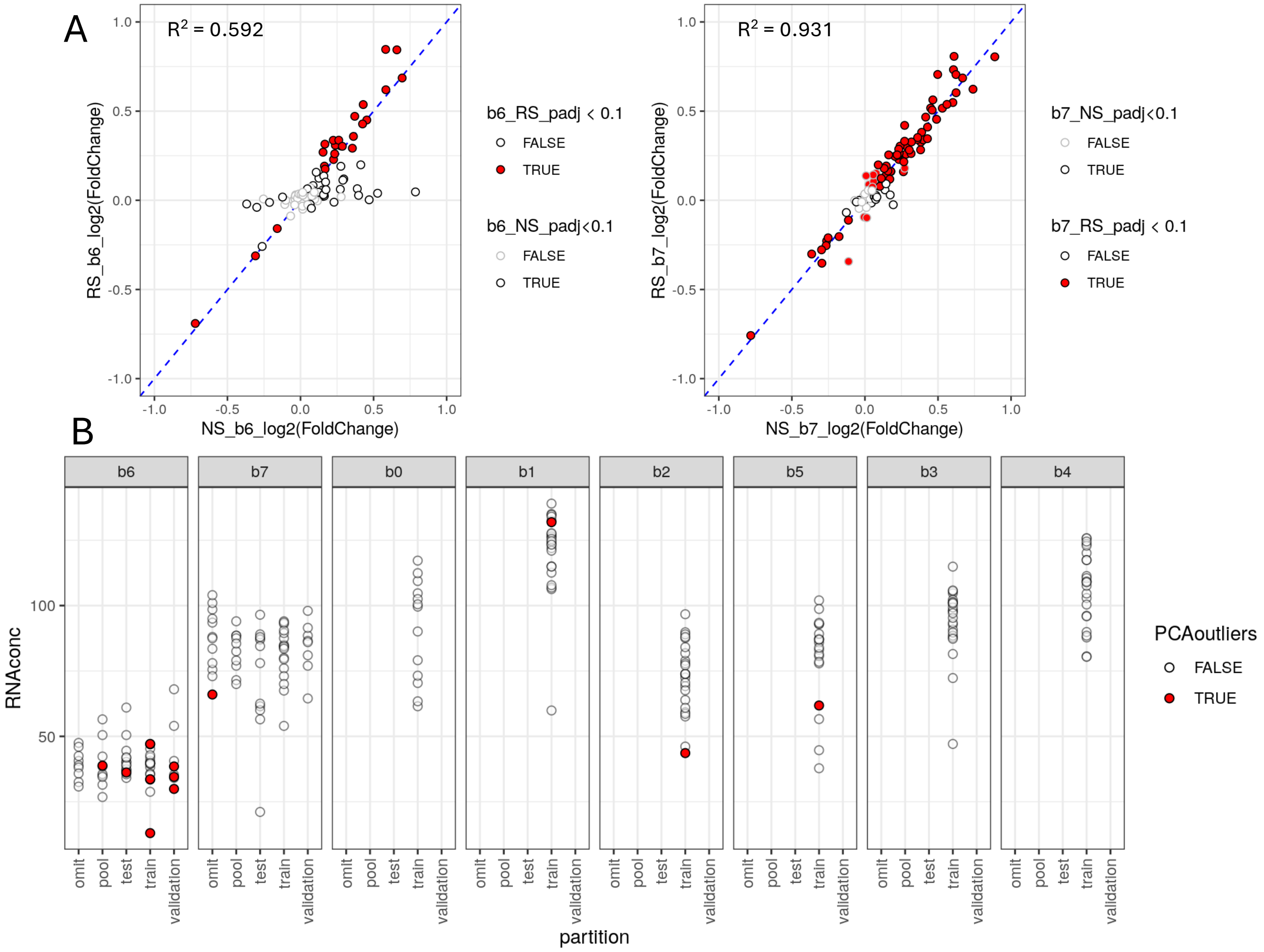


**Figure S3.** Identification of RNAseq failure of LC-iCAP experimental batch 6 by analysis of pooled serum controls and patient samples. *A*, LC-iCAP batches 6 and 7 were each run with 5 technical replicates of case and control pooled serum control pairs and analyzed in separate RNAseq batches. To assess RNAseq data quality, RNA from the pooled controls from batches 6 and 7 were reanalyzed by Nanostring using the development gene set (Data file 2), and differential expression was compared between the Nanostring data (NS) and the RNAseq (RS) data. For batch 7 most genes fell along the identity line (*right*), but for batch 6 several genes deviated from the identity line (*left*), suggesting RS failure. These genes were predominantly genes that showed no differential expression in the RS data. Supporting these data, an analysis comparing Pearson correlation of the expression of each gene across samples between platforms showed a median R of 0.3 for batch 6 and median R of 0.8 for batch 7 (*not shown*). *B,* RNA concentrations for patient samples in LC-iCAP Batch 6 (*b6*) are lower than for other batches (*b0-b5, b7*). RNA concentrations for all patient samples processed by LC-iCAP-RNAseq in cohorts 1-3 are plotted. Cohort 1 is in b0, cohort 2 is in b1-b5 and and cohort 3 is b6 and b7. Samples that were identified as outliers by PCA analysis (not shown) are colored *red*. These data show that experimental batch 6 had low RNA concentrations and low RNAseq data quality.

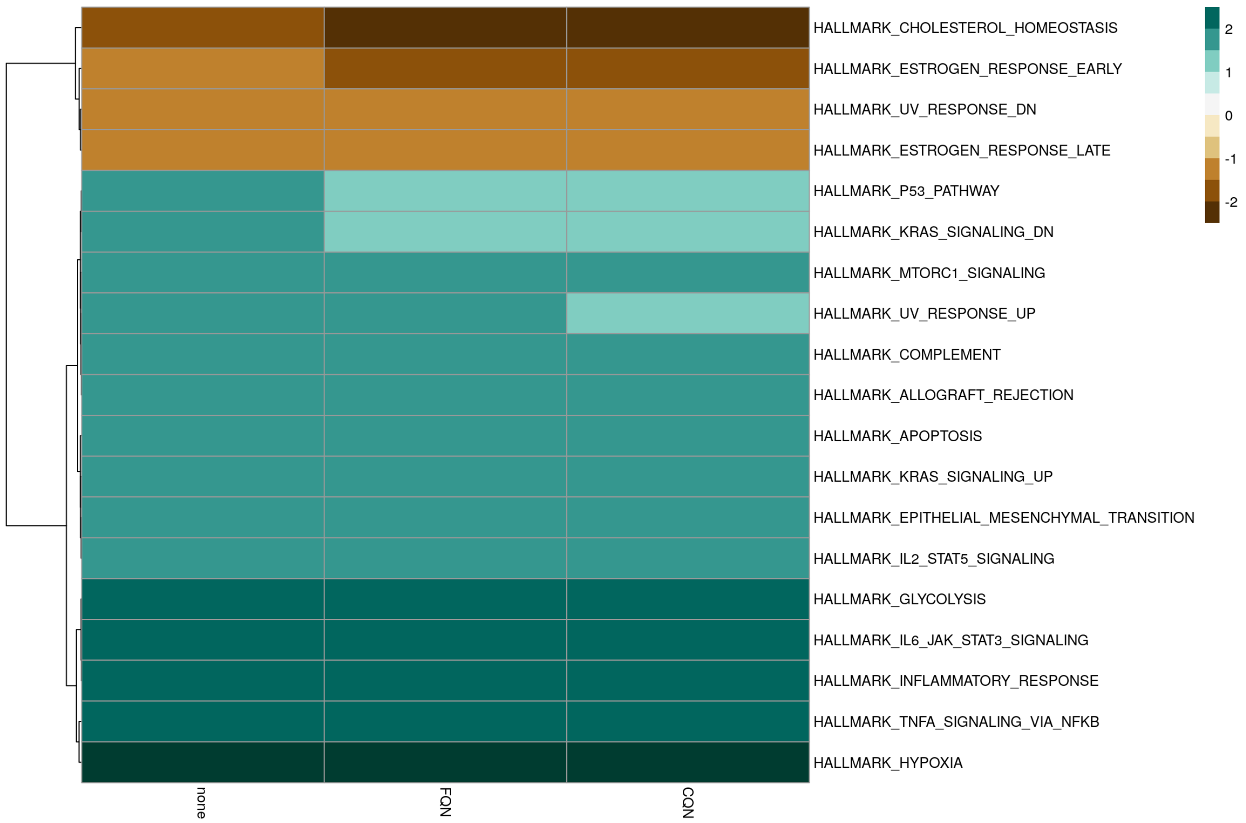


**Figure S4.** Differential response to case versus control serum pools in the LC-iCAP is significantly enriched for HALLMARK functional clusters. Two different experiments with 4 replicates each of the serum pools were merged and used to characterize the response to lung cancer serum by gene set enrichment analysis (GSEA). Significant enrichments were identified for HALLMARK (shown) and oncogenic gene sets (not shown). Only a subset of the MolSigDB gene sets were tested (C1,C3,C4,C6 and H) to avoid set with very large numbers of pathways (C5, C7) and those with restrictions on commercial use (C2). Significantly enriched pathways are those with adjusted p value <0.05 using at least one of the three data processing workflows (without GC bias correction and with FQN or CQN correction.

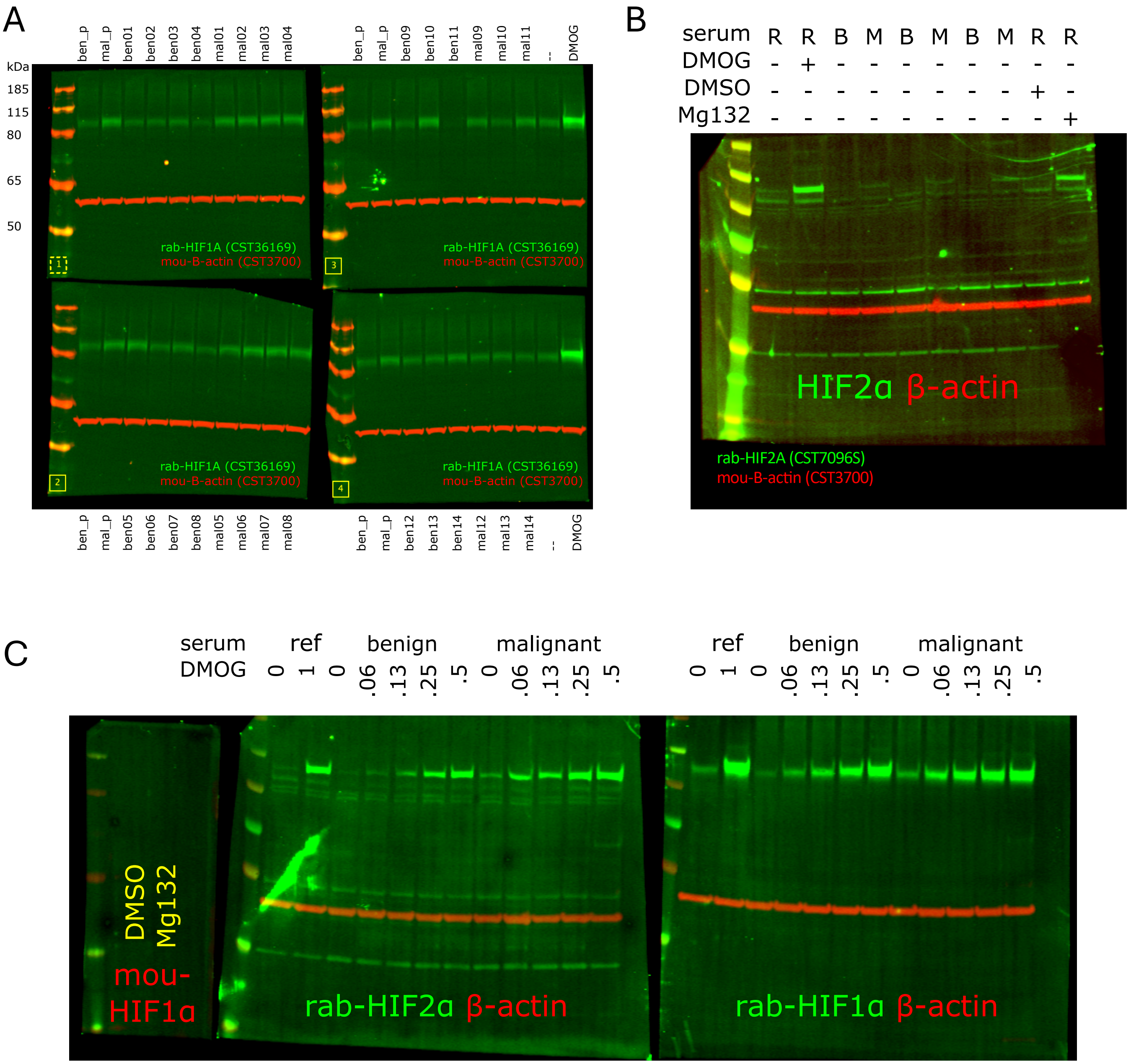


**Figure S5**. Western blot data for graphs in Fig. 5*B left, middle* and *right* are shown in panels *A, B and C.*

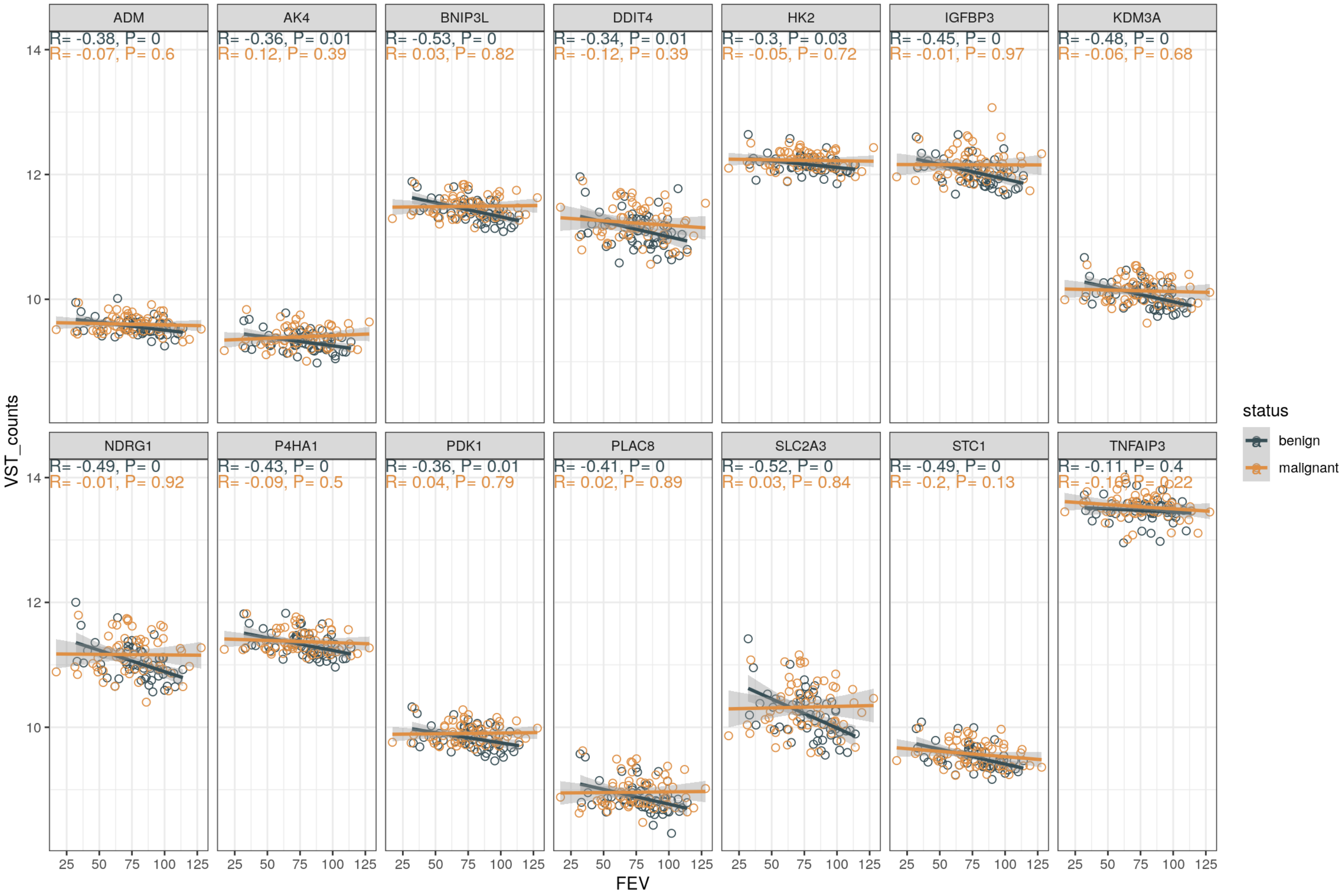


**Fig. S6**. Effect of patient lung function on expression of hypoxia-related genes in the LC-iCAP. For samples in cohorts 1-3, expression values for selected hypoxia-related genes were plotted against patient predicted forced expiratory volume percentage (FEV%) with linear regression lines. For many genes, there is a significant correlation of FEV with gene expression in samples of the benign class suggesting that lung function has a significant effect on the LC-iCAP readout that in some cases affects case versus control differential expression. Pearson correlation coefficients (R) with p values (P) are shown for samples of the benign and malignant classes at the top. For patients with missing values, predicted FEV% was estimated using multivariate linear regression with the following relationship: FEV predicted ~ (pack years + age at collection) * smoking status. The overall R2 was 0.25 with p-value = 0.001. This model was based on a strong linear relationship between FEV% and smoking pack years, the slope of which varied slightly by smoking status.

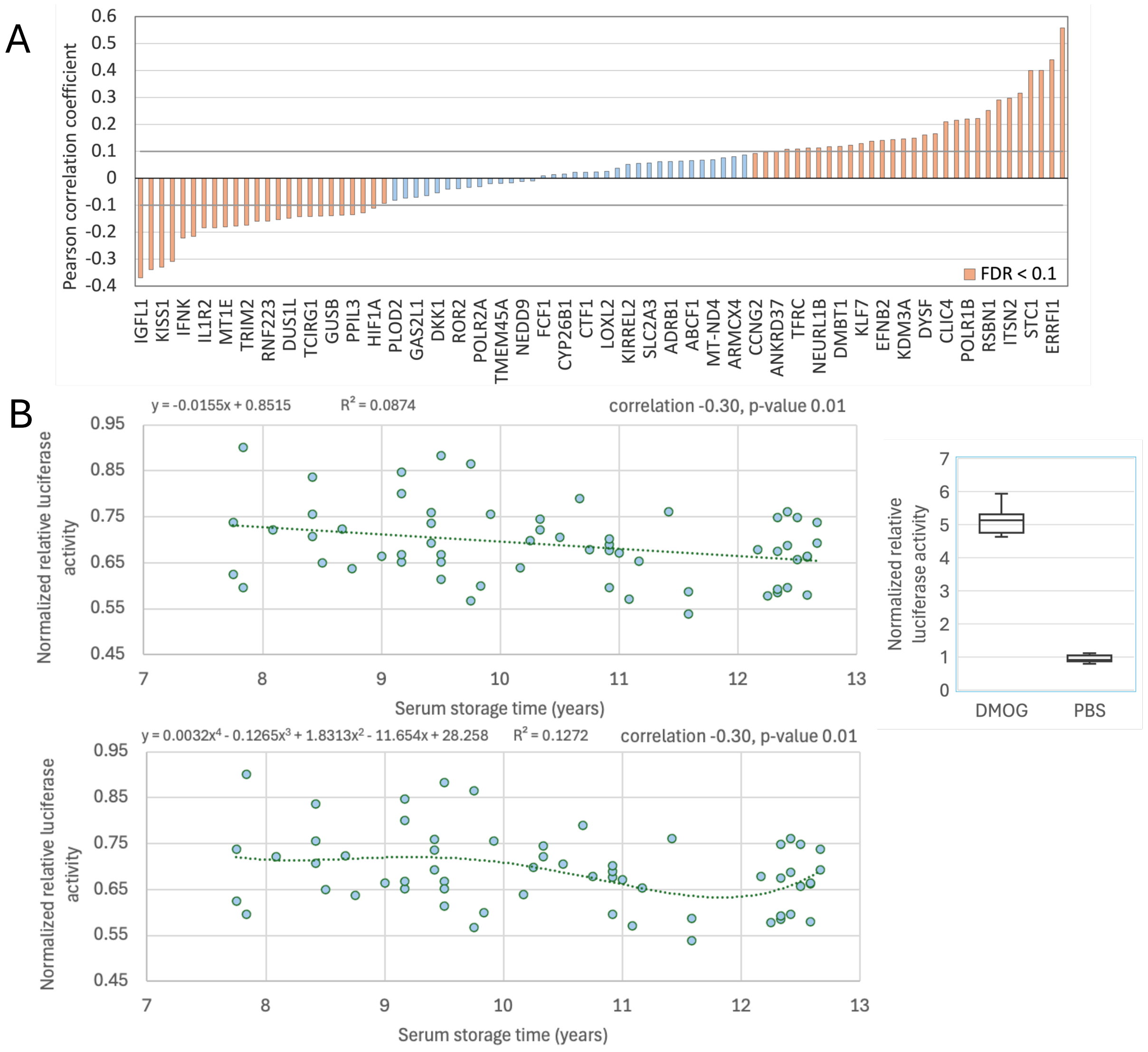


**Figure S7.** Patient serum storage time affects gene expression levels in the LC-iCAP. ***A.*** Bar graphs showing correlation of serum storage time with LC-iCAP gene expression. $Log_2$ gene expression values from LC-iCAP Nanostring Plexset data for 432 samples from cohorts 1-4 (with storage of up to 18 years) were analyzed for correlation with serum storage time. Pearson correlation coefficients (R) for 88 genes with detection above background are sorted in ascending order with names of every other gene labelled. 51 of 88 genes with significant correlation coefficients (FDR < 0.1) are *red*. To control for potential serum source effects, analysis was also conducted with only Vanderbilt (cohorts 1-3 with median storage of 8.6 years) or only University of Pennsylvania samples (cohort 4 with median storage of 13 years) yielding 8 and 35 genes with significant correlation, respectively (data not shown). ***B.*** Scatter plots showing patient serum storage times greater than 10 years have a significant effect on HIF1A activity in the LC-iCAP. A HIF1A-responsive firefly luciferase reporter was stably integrated into 16HBE indicator cells along with a ubiquitously active Renilla luciferase reporter and used to measure HIF1A activity in the LC-iCAP in response to 60 patient serum samples from cohorts 1-2 from Vanderbilt University. Samples were assayed in triplicate using standard LC-iCAP conditions except serum samples had been thawed 3 times. Firefly luciferase activity was averaged across replicates, normalized to Renilla luciferase activity and graphed as a function of serum storage time. Pearson correlation coefficient and p-value are shown indicating significant negative correlation (*upper*). The data were fit to linear regression and a quartic polynomial models in Excel (*upper and lower*). The $R^2$ values were compared, with the quartic model yielding a higher $R^2$ value (0.127 vs. 0.087), indicating a better fit to the data. The quartic model shows a bimodal relationship and indicates better stability of the readout prior to 10 years of storage. These data demonstrate that serum storage time influences the LC-iCAP readout but shows relative stability of the readout for samples stored for less than 10 years. Therefore, a filter was applied during final model development to exclude samples stored for 10 or more years to reduce noise from variable storage times in the assay.

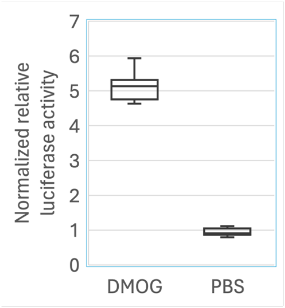


6HRE-TK

CAGAAGTCGACCACGCCGTGTACGTGCTTCACGTGTACGTGGCGATCGTGTACGTGCTGTCTCA**CACAG**CACTCTAG
ACTACTCGTGTACGTGTACGACGTGTACGTGGGCTACGTGTACGTGCTGTCTCA**CACAG**CACGAAGCTTCGTAGCAA
TCGGACCGACGACGttcgc***atattaa***ggtgacgcgtgtggcctcgaacaccgagcgacc*ctgcagcgacccgcttaA*
GGATCCCTGA

**Figure S8** Development of LC-iCAP HIF1A-responsive firefly luciferase reporter assay

To develop a HIF1A-responsive firefly luciferase reporter system used in Figure S8, we constructed and stably integrated a firefly luciferase (ffLUC) reporter into 16HBE indicator cells, along with a ubiquitously active Renilla luciferase reporter for normalization using a lentiviral system. To make the ffLUC reporter, we first made pLV-6HRE-TK-ffLUC, a lentiviral vector with ffLUC under the control of 6HRE-TK consisting of a minimal HSV TK promoter and six copies of the HIF1A-binding hypoxia response elements (6HRE). First, 6HRE-TK was designed based on the sequence of a previously validated 6HRE construct[34]. Next, 6HRE-TK was synthesized and verified by Integrated DNA Technologies (IDT) and delivered in a plasmid (pIDTSmart(Amp)), flanked by SalI and BamHI sites, with an internal HindIII and a site for a Tkrev sequencing primer (Fig. S9*C*). Next, 6HRE-TK was cloned into a commercially available lentiviral vector lacking a promoter (LVR-1048-pLV-Promoterless-Firefly luciferase-PGK-puro, Cellomics Technology) upstream of the ffLUC sequence using SalI and BamHI sites and verified by sequencing. Next, pLV-6HRE-TK-ffLUC was transfected into 293T cells using the EZ-LentiPACK Lentivirus packaging kit (Cellomics Technology) to produce VSV-G pseudotyped lentiviral particles. Next, 16HBE cells were transduced with the produced lentiviral particles (pLV-6HRE TK-fLUC) and a control lentiviral vector expressing Renilla luciferase (cmv-renilla-luciferase-lentivirus, Cellomics Technology). Following transduction, cells were selected using Neomycin and Puromycin to establish HRE-LUC, a stable cell line expressing both luciferase reporters. The response of the HIF1A luciferase reporter to positive and negative controls, DMOG and PBS, is shown ($p$ value < 1E-10) (*upper*). The sequence of 6HRE-TK used to construct the HIF1A-responsive firefly luciferase reporter strain is also shown (*lower*). Restriction sites are underlined; 6 HRE elements are highlighted in gray, induction elements are in blue font; the minimal human HSV TK promoter is small case; the position of TKrev sequencing primer is italicized; TATA box is bold and Italicized.

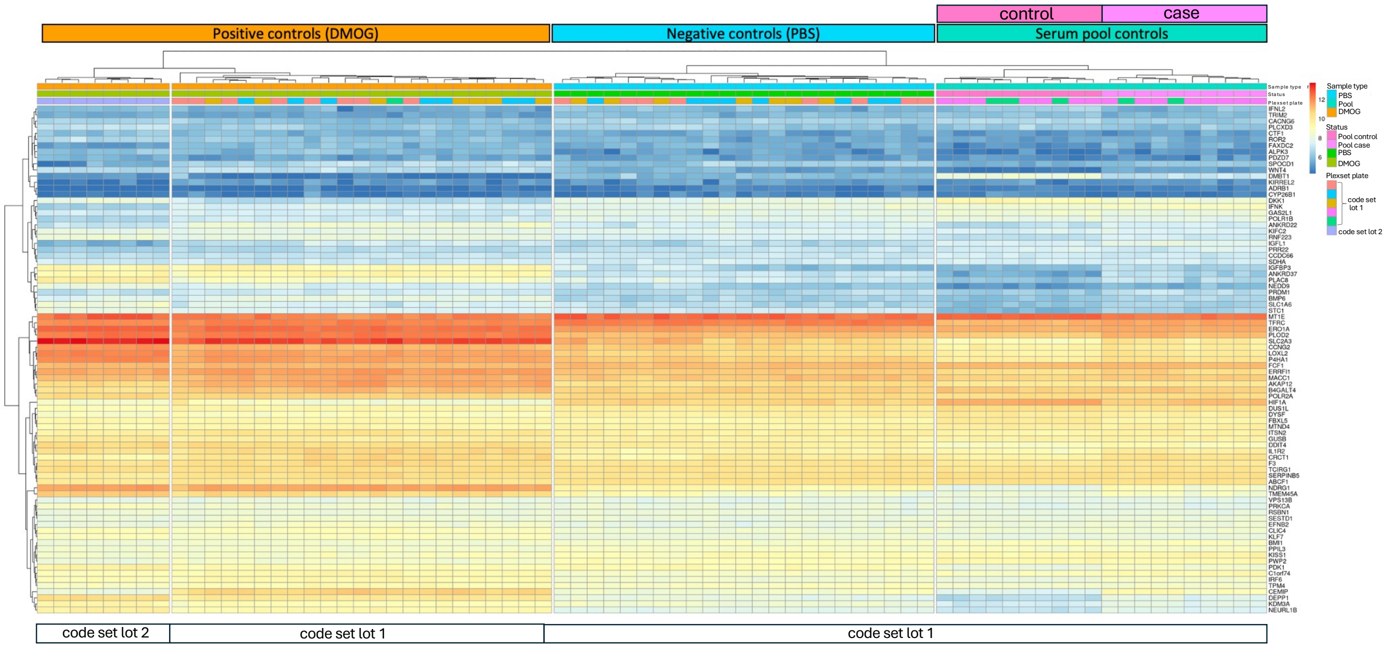


**Figure S9.** LC-iCAP gene expression is reproducible across different LC-iCAP batches, NanoString Plexset plates and across different users. NanoString Plexset data for the 74 standard control samples run with patient samples were analyzed by unsupervised hierarchical clustering (HCL) to measure assay reproducibility. Log2 gene expression values are represented by a two-color gradient. Colored bars at the top of the graph show the control sample type and Plexset plate for each sample and white bars at the bottom indicate code set lot numbers. Dendograms at the top and left side of the graph show relationships of conditions and genes, respectively. Samples partitioned into clusters by sample type and status rather than LC-iCAP batch or Plexset plate suggesting that differential gene expression can be detected above background of batch effects from LC-iCAP batch (across several months), Plexset plate, and different users. However, a code set lot effect was identified based the DMOG samples (left): samples using code set lot 1 clustered separately from those using code set lot 2. These data indicate the presence of code set lot miscalibration, which results in a training set versus blind test set batch effect for 50 genes (see Methods). A successful approach to mitigate this miscalibration was to omit the 50 affected genes from the final model development.

| Model | Test AUC | Algorithm |
|---|---|---|
| M15 | 0.61 (0.48-0.74) | GLM |
| M15R2 | 0.6 (0.48-0.73) | GLM |
| M3 | 0.64 (0.51-0.76) | RF |
| M4 | 0.62 (0.5-0.75) | RF |

**Figure S10.** Model development with LC-iCAP NanoString Plexset data yielded two models with significant performance in blind validation. To address the code set lot miscalibration (identified in Figure S9), we filtered out the 50 most measurably mis-calibrated genes before model development (Data file 4). 4 models were tested on the blind sample set using either generalized linear models (GLM) or random forest (RF) algorithms. The two RF models (M3 and M4) achieved statistically significant performance, highlighting the superiority of the RF ensemble approach in overcoming disease heterogeneity and residual effects of mis-calibration. M3 and M4 are described in Figs. 7 and S11. For comparison, a total of five models were trained and tested on the blind set prior to exclusion of the 50 genes and did not have significant performance.

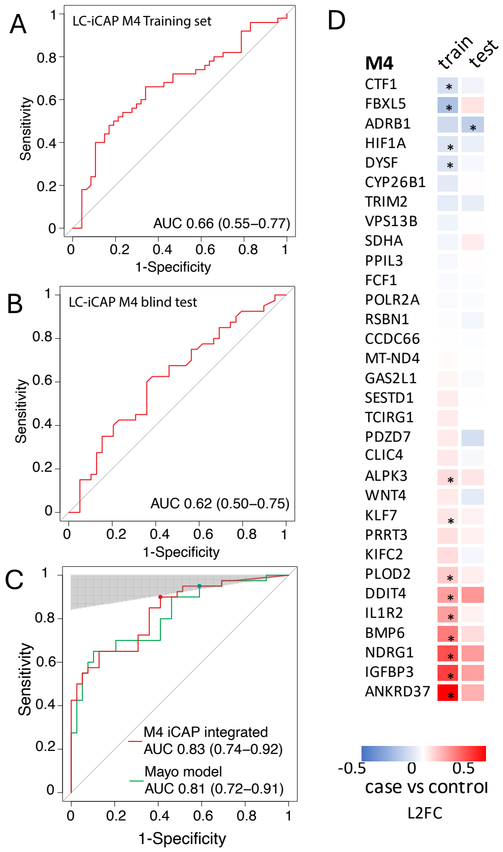


**Figure S11.** ROC curves showing performance of the M4 LC-iCAP model and the M4 iCAP-integrated classifier in temporal blind validation on the test set. ***A and B.*** The M4 LC-iCAP model demonstrates significant performance based on out-of-bag (OOB) predictions from the training set (A) and on an independent blind test set (B) ***C.*** ROC curves showing that the iCAP-integrated classifier outperforms the Mayo Clinic model on the test set. The *shaded* region corresponds to ≥ 95% NPV using a 25% prevalence estimated in the clinical population. The model performances were compared at cut points with clinical utility as rule-out tests (with ≥ 95% NPV and ≥ 90% sensitivity) (*colored nodes*), and the integrated classifier had significantly higher specificity than the Mayo model (exact binomial test p-value = 0.016). The cut points correspond to 96.1% NPV, 41% specificity and 95% sensitivity for the Mayo model and 94.7% NPV, 59% specificity and 90% sensitivity for the iCAP integrated classifier. *AUC*, area under the receiver operating characteristic (*ROC*) curve are shown with 95% confidence intervals in *brackets* (DeLong); *NPV*, negative predictive value. ***D.*** Heat map of LC-iCAP NanoString Plexset data showing case versus control differential expression for genes of the M4 model. For samples of the training and test sets, median differential expression values are shown for genes detected above background in >50% of samples. Genes with significant differential expression are indicated with asterisks (Mann Whitney U test p-values <0.05). Training versus test set differential expression profiles had a strong Pearson correlation coefficient of 0.73 (*p*-value 2.5E-6) showing good reproducibility of expression between training and blind test sets. L2FC, $\log_2$ fold change.

$$iCAP\ I(k) = \begin{cases} \max(0, p(k) - 0.4), & LC\ iCAP\ probability\ of\ malignancy \leq 0.49 \\ p(k), & LC\ iCAP\ probability\ of\ malignancy > 0.49 \end{cases}$$

$$p(k) = \frac{e^X}{1 + e^X} \qquad X = -6.8272 + (0.0391 * Age) + (0.7917 * Smoker) + (1.3388 * \text{cancer}) + (0.1274 * Diameter) + (1.0407 * Spiculation) + (0.7838 * Location)$$

**Figure S12.** Technical definition and equation for the iCAP integrated classifier using M3. The classifier integrates the probability of malignancy from the LC-iCAP model with that from the Mayo model (*p(k)*) based on 5 clinical risk factors (Swensen, 1997). The integrated classifier provides a numerical value *iCAP I* for a subject *k*, as defined above: where *Age* is the age of the subject in years, *Smoker* is 1 if the subject is a former or current smoker (otherwise 0), Cancer is 1 if the patient has a history of cancer, 0 if not. *Diameter* is the size of the lung nodule in mm, *Spiculation* is 1 if the lung nodule is spiculated (otherwise 0) and *Location* is 1 if the lung nodule is located in an upper lung lobe (otherwise 0). *iCAP I(k)* ranges between 0 and 1; LC iCAP probability of malignancy scores above and below 0.49 are positive and negative for malignancy, respectively. For the M4 integrated classifier, the LC iCAP probability of malignancy threshold was 0.407.
